# Transparency in infectious disease research: a meta-research survey of specialty journals

**DOI:** 10.1101/2022.11.11.22282231

**Authors:** Emmanuel A. Zavalis, Despina G. Contopoulos-Ioannidis, John P.A. Ioannidis

## Abstract

**Introduction:** Infectious diseases carry a large global burden and have implications for society at large. Therefore, reproducible, transparent research is extremely important. To assess the current state of transparency in this field, we investigated code sharing, data sharing, protocol registration, conflict of interest and funding disclosures in articles published in the most influential infectious disease journals.

**Methods:** We evaluated transparency indicators in the 5340 PubMed Central Open Access (PMC OA) articles published in 2019 or 2021 in the 9 most-cited specialty journals in infectious disease. We used a previously validated text-mining R package, *rtransparent*. The approach was manually validated for a random sample of 200 articles for which study characteristics were also extracted in detail. Main comparisons assessed 2019 versus 2021 articles, 2019 versus 2021 non-COVID-19 articles, and 2021 non-COVID-19 articles versus 2021 COVID-19 articles.

**Results:** A total of 5340 articles were evaluated (1860 published in 2019 and 3480 in 2021 (of which 1828 on COVID-19)). Text-mining identified code sharing in 98 (2%) articles, data sharing in 498 (9%), registration in 446 (8%), conflict of interest disclosures in 4209 (79%) and funding disclosures in 4866 (91%). There were substantial differences across the 9 journals in the proportion of articles fulfilling each transparency indicator: 1-9% for code sharing, 5-25% for data sharing, 1-31% for registration, 7-100% for conflicts of interest, and 65-100% for funding disclosures. There were no major differences between articles published in 2019 and non-COVID-19 articles in 2021. In 2021, non-COVID-19 articles had more data sharing (12%) than COVID-19 articles (4%). Validation-corrected imputed estimates were 3% for code sharing, 11% for data sharing, 8% for registrations, 79% for conflict of interest disclosures and 92% for funding disclosures.

**Conclusion:** Data sharing, code sharing, and registration are very uncommon in infectious disease specialty journals. Increased transparency is required.

## 1 Introduction

Infectious diseases are an important field in medicine, epidemiology, and public health across a spectrum that spans basic science, translational research, clinical and population-level applications. The global burden of infectious disease has been large, with a higher share in less developed countries(1–4). With the advent of the COVID-19 pandemic the developed world was sensitized to the field with awakened interest and its funding and research output increased rapidly(5–7). The rigor and reliability of the evidence generated in the field of infectious diseases therefore has major implications for the health of individuals, populations, and societies at large. In this regard, transparency features, such as sharing of data and code, availability of pre-registered protocols, as well as reporting of conflicts of interest and funding, can be fundamental in evaluating the evidence obtained by research investigations in infectious diseases (8–10). Previous work has assessed these transparency indicators in depth in infectious disease models specifically, a type of research that became highly popular and influential during the COVID-19 pandemic(11). It was found that the majority of such epidemiological modeling studies do not share their data and/or their code, and very few have registered protocols. The vast majority of published articles have conflict of interest and funding statements, but it is not clear whether the information reported is complete(12,13).

Infectious disease research, nevertheless, encompasses a very large range of study designs and research efforts. It is unknown whether transparency is high for these diverse types of designs and whether there are some study features and characteristics that may be associated with better or worse performance of the published articles in terms of transparency indicators. More importantly, the COVID-19 pandemic was a crash test for many scientific fields, and most prominently this applied par excellence to the field of infectious diseases. Massive publication volume may not necessarily have been accompanied by high quality and/or transparency(5,14,15). It is important to study whether the infectious diseases literature published during the pandemic was different in this regard compared to the pre-pandemic papers published in the same journals that specialize on infectious diseases.

Here, we present the results of meta-epidemiological assessment of transparency indicators of recent articles published in the major infectious disease specialty journals, comparing the transparency performance of these journals in the pre-pandemic (2019) and in the COVID-19 pandemic (2021) period.

## 2 Materials and Methods

The protocol of the study was pre-registered in Open Science Framework (https://doi.org/10.17605/OSF.IO/FYZPX).

### 2.1 Study sample

We examined the papers published in 2019 and 2021 by the 9 specialty journals that received the largest total number of citations in infectious disease research, according to the InCites Journal Citation Reports (JCR)(16) in the respective specialty category (see Supplementary Table 1). We focused on papers indexed in the PubMed Central Open Access (PMC OA) subset of PubMed, similar to previous work, since these are the papers that can be massively downloaded for in depth text mining of transparency indicators. The specific journals are tabulated in Supplementary Table 1 along with the number of citations that they received based on the latest JCR edition(16). We aimed to focus on original articles and reviews, excluding letters, editorials, and study protocols.

Supplementary Table 1

### 2.2 Search query

The search was performed on PubMed on the 14^th^ of September 2022 using the query: “pubmed pmc open access”[Filter] AND (“clin infect dis”[Journal] OR “j infect dis”[Journal] OR “lancet infect dis”[Journal] OR “infect immun”[Journal] OR “emerg infect dis”[Journal] OR “j antimicrob chemother”[Journal] OR “clin microbiol infect”[Journal] OR “bmc infect dis”[Journal] OR “int j infect dis”[Journal]) AND (“journal article”[Publication Type]) NOT (“editorial”[Publication Type] OR “letter”[Publication Type] OR “clinical trial protocol”[Publication Type]) AND “hasabstract”[All Fields] AND “english”[Language] AND (ffrft[Filter]) AND (2019:2019[pdat] OR 2021:2021[pdat])

### 2.3 Data extraction

#### 2.3.1 Text mining

For each eligible article we used PubMed to extract information on metadata that included PMID, PMCID, publication year, journal name, affiliation and the R package *rtransparent*(17) to extract the following transparency indicators: (i) code sharing (ii) data sharing (iii) protocol (pre-)registration, (iv) conflicts of interest and (v) funding statements. *rtransparent* was also used to retrieve whether a journal was original research or a review, so as to be eligible. *rtransparent* searches through the full text of the papers for specific words or phrases that strongly suggest that the aforementioned transparency indicators are present in that particular paper. The program uses regular expressions to adjust for variations in expressions. For details on the exact code and terms/phrases captured by *rtransparent*, see(17).

#### 2.3.2 Manually extracted characteristics

From a random sample of 200 articles (2019 and 2021) we extracted additional characteristics manually. The manually extracted characteristics were meta-data (country of the first and last author, length of the article, number of tables, figures and appendices); study design; type of pathogen/ disease/ drug or vaccine studied; and, for clinical studies, whether a study was interventional or not, and its primary focus if interventional (therapy or prevention). Finally, we extracted information about the study population including age group, human vs animal work and sample size (for systematic reviews we used the number of included articles). See our protocol of our study (https://doi.org/10.17605/OSF.IO/FYZPX) for details on items extracted and options for each item.

### 2.4 Primary comparisons and statistical analysis

We considered three primary comparisons that were conducted using Fisher’s exact tests for each transparency indicator separately. For analysis of associations, we used a statistical significance threshold of 0.005(18). The analysis code was written in R 4.2.1(19).

We compared transparency indicators for 2019 versus 2021, 2019 versus 2021 non-COVID-19, and 2021 COVID-19 versus 2021 non-COVID-19 articles in order to evaluate whether there has been an improvement over time, and whether COVID-19 articles differed from non-COVID-19 ones. These comparisons were performed with and without stratification for journal, since it was possible that differences are seen across specific journals. Fisher’s exact tests were used to test our unstratified comparisons. The tests were performed for each transparency indicator separately. To stratify and account for journals we had originally anticipated to perform random-effects meta-analyses for odds ratios, stratifying the overall sample by journal. However, there was no significant between-journal heterogeneity for almost all analyses (with rare exceptions), and some cells in 2×2 tables for some journals had very small numbers and zeros. Therefore, we preferred as the default the fixed effects analysis with a 0.5 correction for 2×2 tables with zero cells, but random effects are also presented.

The COVID-19 articles were identified by categorizing all retrieved articles when combining the main search query to the following query in PubMed: SARS-CoV-2 OR ‘coronavirus 2’ OR ‘corona virus 2’ OR covid-19 OR {novel coronavirus} OR {novel corona virus} OR 2019-ncov OR covid OR covid19 OR ncovid-19 OR ‘coronavirus disease 2019’ OR ‘corona virus disease 2019’ OR corona-19 OR SARS-nCoV OR ncov-2019 used in prior research(20).

### 2.5 Secondary comparisons

Secondary comparisons of the transparency across the manually extracted characteristics were performed. The comparisons were descriptive rather than testing specific hypotheses and they focused on code sharing, data sharing and registration since it was expected (and indeed documented eventually) that the vast majority of articles would have a place-holder statement for conflict of interest and funding.

### 2.6 Manual validation

In order to validate the performance of the automated text mining algorithms, we evaluated the 200 randomly selected articles manually for the presence of each of the 5 text mined indicators. This aimed to identify for each indicator, the number of false positives (indicator identified in text mining, but not manually) and false negatives (indicator not identified in text mining but identified manually).

This information allowed a correction of the estimates of the proportion of articles that satisfy each of the transparency indicators. As in previous work(11), the corrected proportion C(i) of publications satisfying an indicator i was obtained by U(i) × TP + (1 − U(i)) × FN, where U(i) is the uncorrected proportion detected by the automated algorithm, TP is the proportion of true positives (proportion of those manually verified to satisfy the indicator among those identified by the algorithm as satisfying the indicator), and FN is the proportion of false negatives (proportion of those manually found to satisfy the indicator among those categorized by the algorithm not to satisfy the indicator).

In the 200 randomly selected articles, moreover, whenever there was a conflict of interest disclosure statement, we noted whether no conflict was disclosed (e.g. “the authors have no conflicts to disclose”. Similarly, for funding, we noted whether the statement stated that no funding was received, or specific funding was disclosed.

### 2.7 Amendments to the original protocol

1. During the manual validation procedure, we noted that a number of articles in Clinical Infectious Diseases and in Journal of Infectious Diseases were available in PMC only in a watermarked pdf version that could not be accessed by the automated algorithm. We therefore went back and identified all 293 articles in our sample that could not be read by the automated algorithms and manually extracted them for the 5 transparency indicators.
2. Fixed effects were preferred over random effects for the journal-stratified analyses, as discussed above. They practically give identical results when the estimated between-study variance is 0 (in the vast majority of cases). Fixed effects is preferable given the very small numbers in some 2×2 table cells and acceptable given the lack of between-study heterogeneity (with few exceptions, as discussed below). Random effects estimates are also provided.

## 3 Results

### 3.1 Overall study sample

The search query retrieved 5701 articles from the PubMed Central Open Access subset of which 361 were neither an original article nor a systematic review (using *rtransparent* classification of articles) and were excluded. Of the 5340 eligible papers, 1860 were published in 2019 and 3480 in 2021. 19 papers published in 2019 were retrieved by the COVID-19 search query, but upon in depth examination none proved to be COVID-19 research but reflected a 1% false COVID-19 assignment error. 1828/3480 articles published in 2021were retrieved with the COVID-19 query (See Figure 1).

**Figure 1.**
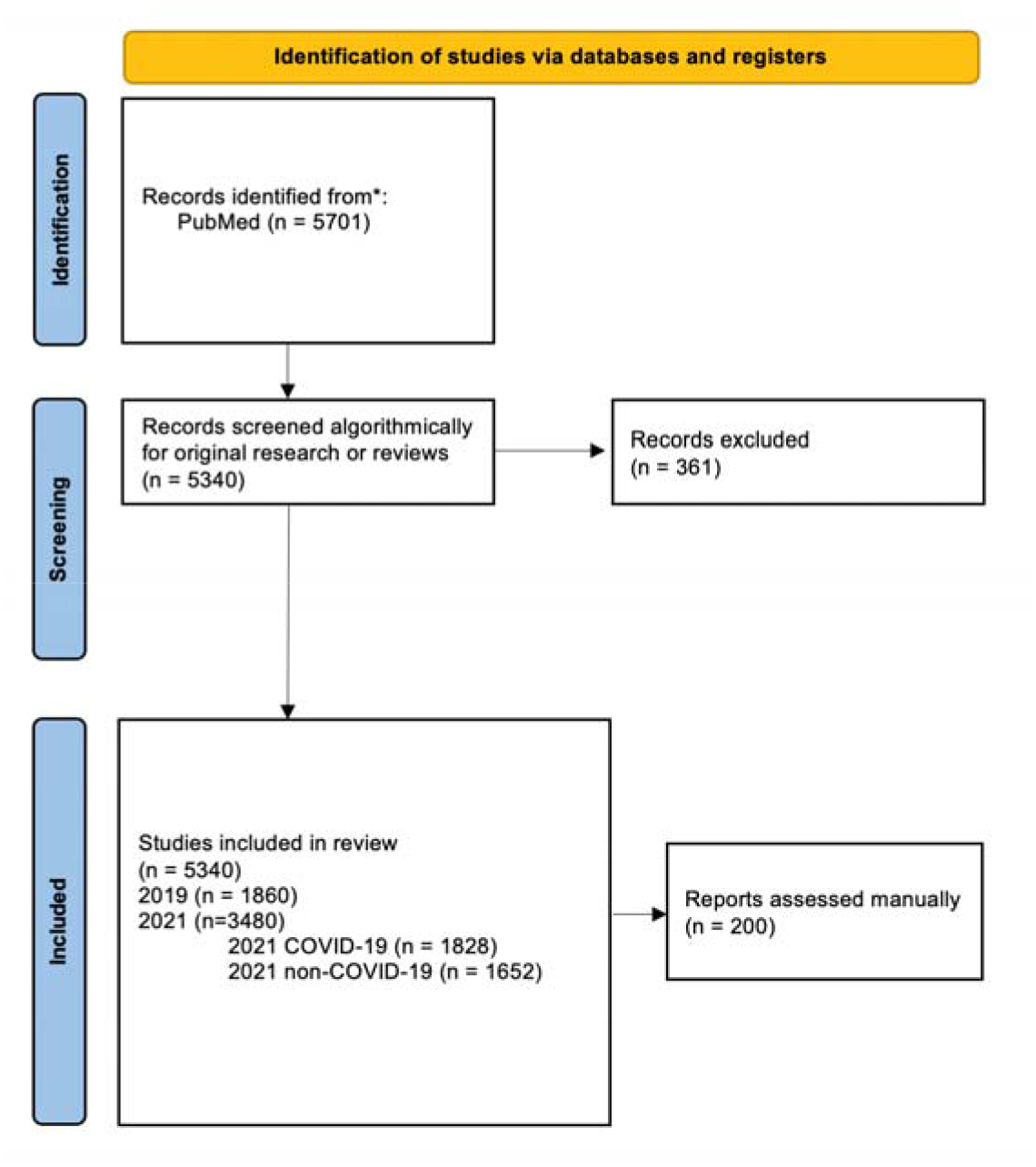
Study selection flowchart

Of the 5340 publications, there were 1956 (37%) from BMC Infectious Diseases, 850 (16%) from Clinical Infectious Diseases, 194 (4%) from Clinical Microbiology and Infection, 912 (17%) from Emerging Infectious Diseases, 107 (2%) from Infection and Immunity, 585 (11%) from International Journal of Infectious Diseases, 212 (4%) from Journal of Antimicrobial Chemotherapy, 383 (7%) from Journal of Infectious Diseases, and 141(3%) from Lancet Infectious Diseases. The proportion of COVID-19 publications in 2021 ranged from 0% for Infection and Immunity to 88% for International Journal of Infectious Diseases.

### 3.2 Transparency indicators

In the examined sample of 5340 articles, 98 (2%) shared code, 498 (9%) shared data, 446 (8%) were registered, 4209 (79%) contained a conflict of interest statement, and 4866 (91%) a funding statement.

When stratifying the transparency indicators across journal there were observable differences across publications in the different journals with some outliers. For instance, Lancet Infectious Diseases had 9% articles that shared code whilst the others had rates around 2%. For data sharing, Infection and Immunity was an outlier with 25% data sharing articles, while Emerging Infectious Diseases and Journal of Antimicrobial Chemotherapy were at 14-15% and all other journals just had 5-10% data sharing. There was also a clear difference in registration rates, with the highest rates in Lancet Infectious Diseases (31%) and the lowest in Infection and Immunity (1%). Rates of conflict of interest disclosures varied widely from 7% in Emerging Infectious Diseases to 100% in BMC Infectious Diseases. Funding disclosures varied modestly, from 65% in Emerging Infectious Diseases to 100% in BMC Infectious Diseases (see Table 1).

**Table 1.**
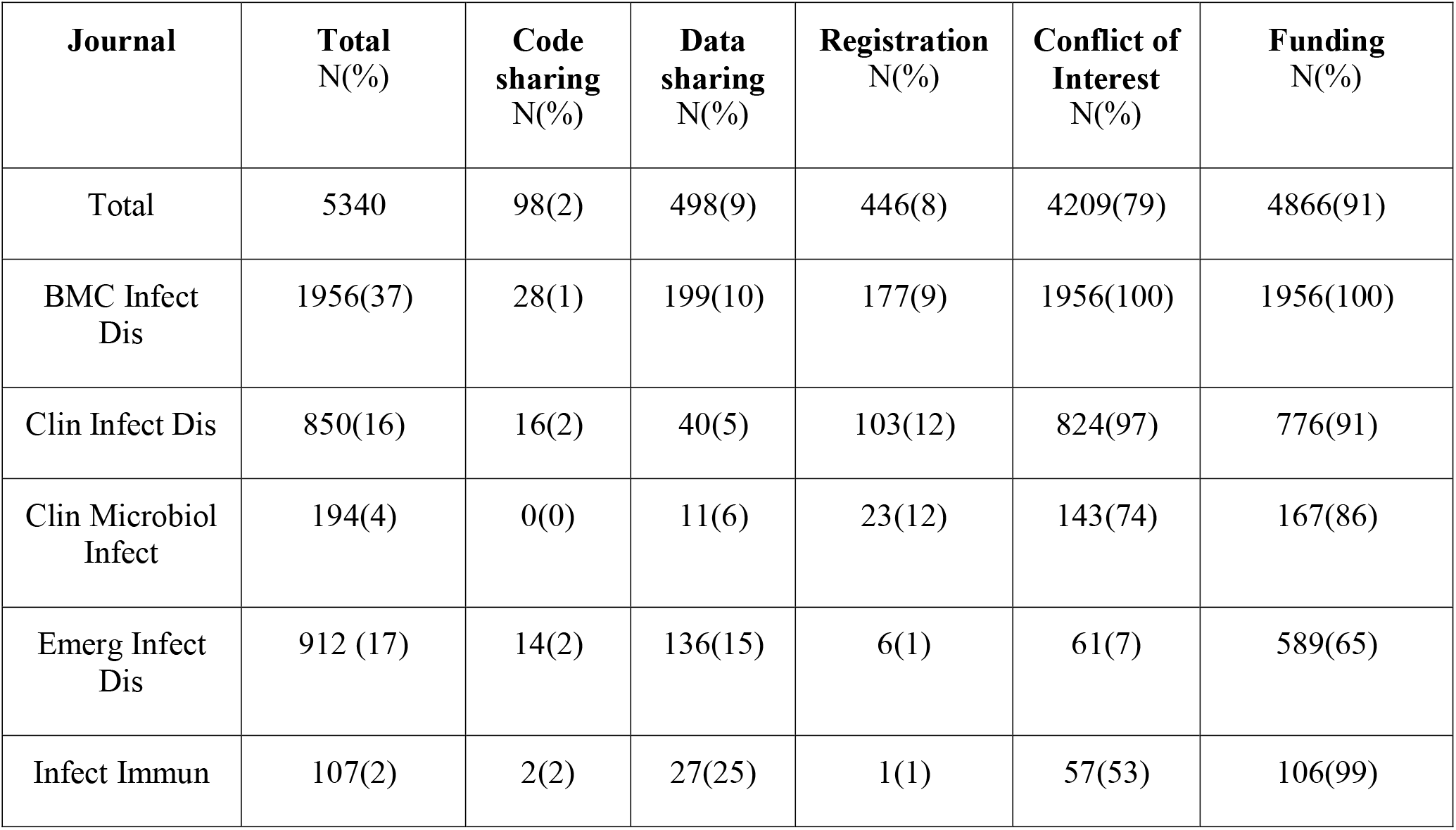
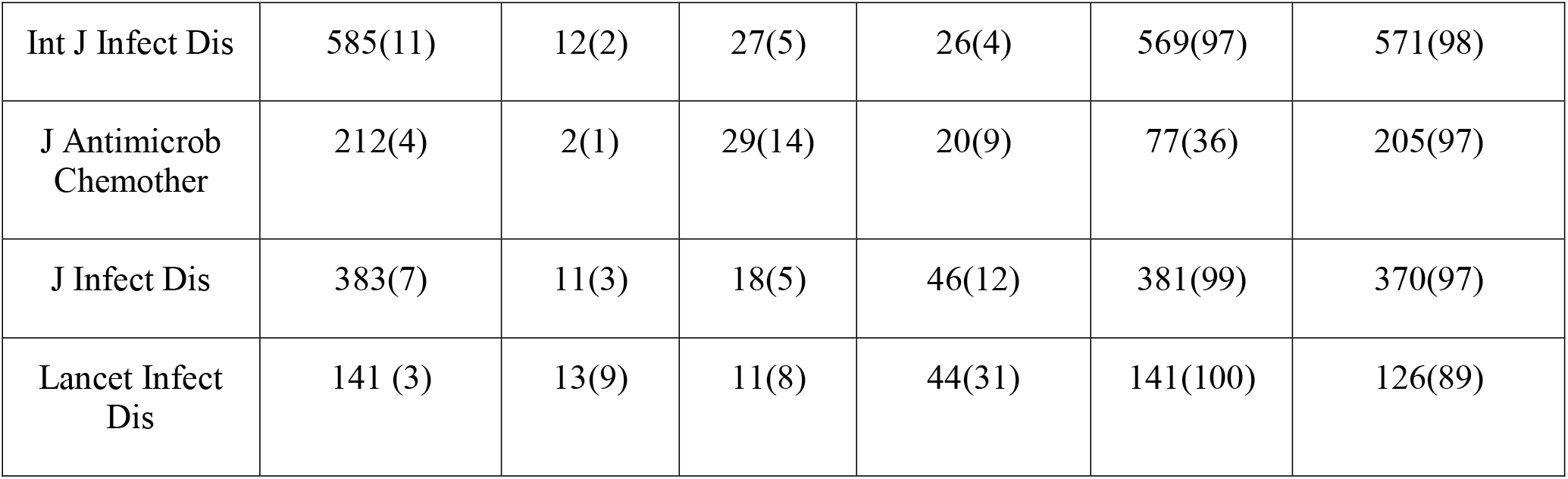
Transparency indicators stratified across journal

| Journal | Total<br>N(%) | Code<br>sharing<br>N(%) | Data<br>sharing<br>N(%) | Registration<br>N(%) | Conflict of<br>Interest<br>N(%) | Funding<br>N(%) |
| --- | --- | --- | --- | --- | --- | --- |
| Total | 5340 | 98(2) | 498(9) | 446(8) | 4209(79) | 4866(91) |
| BMC Infect<br>Dis | 1956(37) | 28(1) | 199(10) | 177(9) | 1956(100) | 1956(100) |
| Clin Infect Dis | 850(16) | 16(2) | 40(5) | 103(12) | 824(97) | 776(91) |
| Clin Microbiol<br>Infect | 194(4) | 0(0) | 11(6) | 23(12) | 143(74) | 167(86) |
| Emerg Infect<br>Dis | 912 (17) | 14(2) | 136(15) | 6(1) | 61(7) | 589(65) |
| Infect Immun | 107(2) | 2(2) | 27(25) | 1(1) | 57(53) | 106(99) |
| Int J Infect Dis | 585(11) | 12(2) | 27(5) | 26(4) | 569(97) | 571(98) |
| J Antimicrob<br>Chemother | 212(4) | 2(1) | 29(14) | 20(9) | 77(36) | 205(97) |
| J Infect Dis | 383(7) | 11(3) | 18(5) | 46(12) | 381(99) | 370(97) |
| Lancet Infect<br>Dis | 141 (3) | 13(9) | 11(8) | 44(31) | 141(100) | 126(89) |

The primary comparisons of transparency indicators between 2019 and 2021 showed a statistically significant increase in code sharing, a decrease in data sharing, and a decrease in funding disclosures, but the differences were very modest in absolute magnitude (all ≤ 4%). Comparing the samples from 2019 and the non-COVID-19 papers from 2021 yielded similar rates of transparency in these two years across all five transparency indicators. Finally, the comparison of the 2021 non-COVID-19 papers to the COVID-19 papers published in the same year showed a statistically significantly higher rate of data sharing and a lower rate of conflict of interest disclosures in non-COVID-19 papers (see Table 2).

**Table 2.** Primary comparisons of transparency indicators

| **N=5340** | **Total** | **Code sharing**<br>**N(%)** | **Data sharing**<br>**N(%)** | **Registration**<br>**N(%)** | **Conflict of Interest**<br>**N(%)** | **Funding**<br>**N(%)** |
| --- | --- | --- | --- | --- | --- | --- |
| 2019 | 1860 | 19(1) | 219(12) | 168(9) | 1439(77) | 1731(93) |
| 2021 | 3480 | 79(2) | 279(8) | 278(8) | 2770(80) | 3135(90) |
| non-COVID-19 | 1652(37) | 27(2) | 204(12) | 152(9) | 1280(77) | 1507(91) |
| COVID-19 | 1828(53) | 52(3) | 75(4) | 126(7) | 1490(82) | 1628(89) |
| **Fisher's exact test p-values** |  |  |  |  |  |  |
| 2019 vs 2021 | | 0.0009 | $1.45 \times 10^{-5}$ | 0.19 | 0.06 | 0.0002 |
| 2019 vs 2021 non-COVID-19 |  | 0.14 | 0.60 | 0.86 | 0.94 | 0.04 |
| 2021 non-COVID-19 vs 2021 COVID-19 | | 0.02 | $< 2.2 \times 10^{-16}$ | 0.012 | 0.004 | 0.04 |

Meta-analysis of the data stratified per journal showed no statistically significant heterogeneity in any of the comparisons with the exception of funding disclosures (I^2^=72%(44%-86%)) and registration (I^2^=65% (22%-85%)) for the comparison of COVID-19 versus non-COVID-19 articles in 2021. Meta-analytic results were largely similar to those inferred by Fisher’s exact tests without stratifying for journal (see Table 3).

**Table 3.** Meta-analyses of primary comparisons

| Summary | Summary OR(95%CI) by fixed effects | Summary OR(95%CI) by random effects | Heterogeneity I<sup>2</sup> % (95% CI) | Heterogeneity p-value |
| --- | --- | --- | --- | --- |
| Code sharing (2021 v. 2019) | 1.78(1.06-2.98) | 1.78(1.06-2.98) | 0(0-65) | 0.66 |
| Code sharing (2021 non-COVID-19 v. 2019) | 1.62(0.87-3.01) | 1.62(0.87-3.01) | 0(0-65) | 0.7 |
| Code sharing (2021 COVID-19 v. 2021 non-COVID-19) | 1.65(0.98-2.75) | 1.65(0.98-2.75) | 22(0-65) | 0.24 |
| Data sharing (2021 v. 2019) | 0.77(0.64-0.94) | 0.77(0.64-0.94) | 15(0-57) | 0.31 |
| Data sharing (2021 non-COVID-19 v. 2019) | 1.07(0.87-1.31) | 1.10(0.86-1.40) | 0(0-65) | 0.44 |
| Data sharing (2021 COVID-19 v. 2021 non-COVID-19) | 0.36(0.26-0.50) | 0.36(0.24-0.53) | 4(0-66) | 0.40 |
| Registration (2021 v. 2019) | 0.83(0.67-1.03) | 0.82(0.63-1.07) | 0(0-65) | 0.49 |
| Registration (2021 non-COVID-19 v. 2019) | 1.00(0.79-1.28) | 1.01(0.79-1.28) | 0(0-65) | 0.43 |
| Registration (2021 COVID-19 v. 2021 non-COVID-19) | 0.65(0.49-0.87) | 0.78(0.43-1.42) | 72(44-86) | <0.01 |
| COI disclosures (2021 v. 2019) | 1.08 (0.77-1.51) | 1.08(0.77-1.51) | 40(0-75) | 0.13 |
| COI disclosures (2021 non-COVID-19 v. 2019) | 1.16(0.80-1.68) | 1.16(0.80-1.68) | 20(0-83) | 0.29 |
| COI disclosures<br>(2021 COVID-19<br>v. 2021 non-COVID-19) | 0.90(0.56-1.42) | 0.81(0.44-1.49) | 37(0-73) | 0.15 |
| Funding<br>disclosures (2021<br>v. 2019) | 0.67(0.53-0.86) | 0.60(0.36-0.98) | 27(0-67) | 0.21 |
| Funding<br>disclosures (2021<br>non-COVID-19 v.<br>2019) | 0.72(0.54-0.97) | 0.72(0.54-0.97) | 5(0-72) | 0.39 |
| Funding<br>disclosures (2021<br>COVID-19 v. 2021<br>non-COVID-19) | 0.84(0.63-1.11) | 0.65(0.32-1.32) | 65(22-85) | <0.01 |

In Table 4 we have tabulated the transparency according to the groups of the main comparisons and journal and performed stratified Fisher’s exact tests for all hypotheses and all journals separately for each indicator.

**Table 4.**
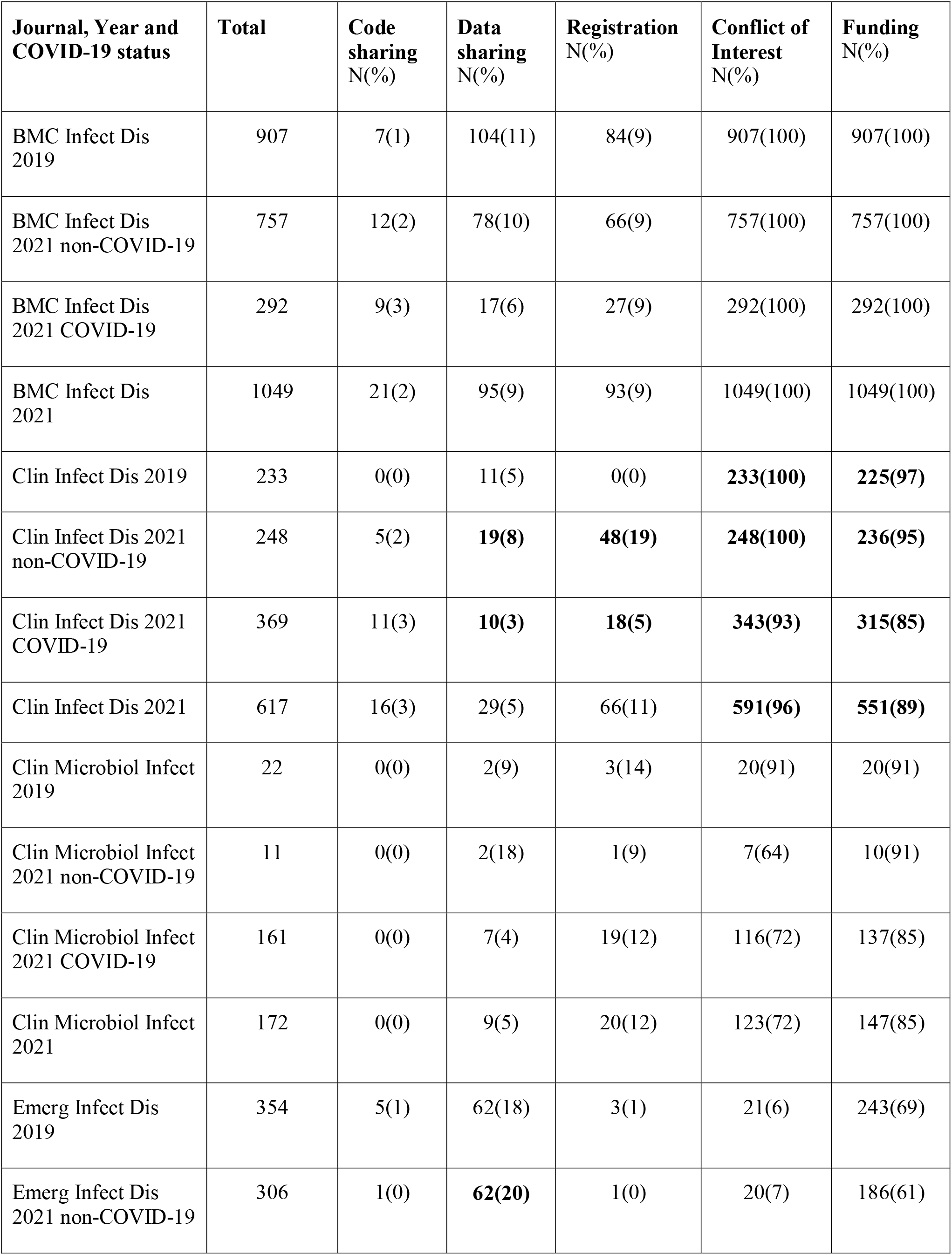
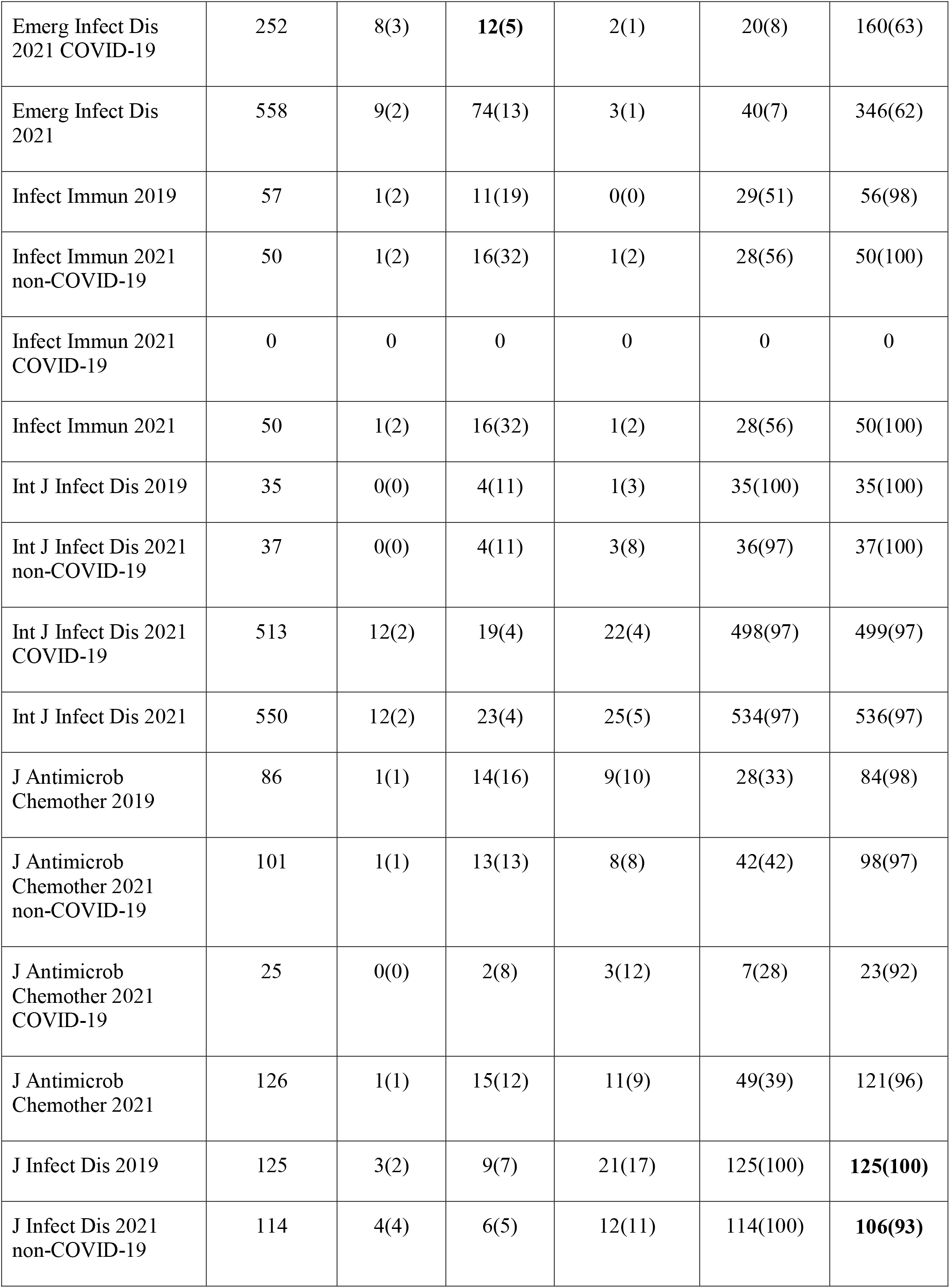
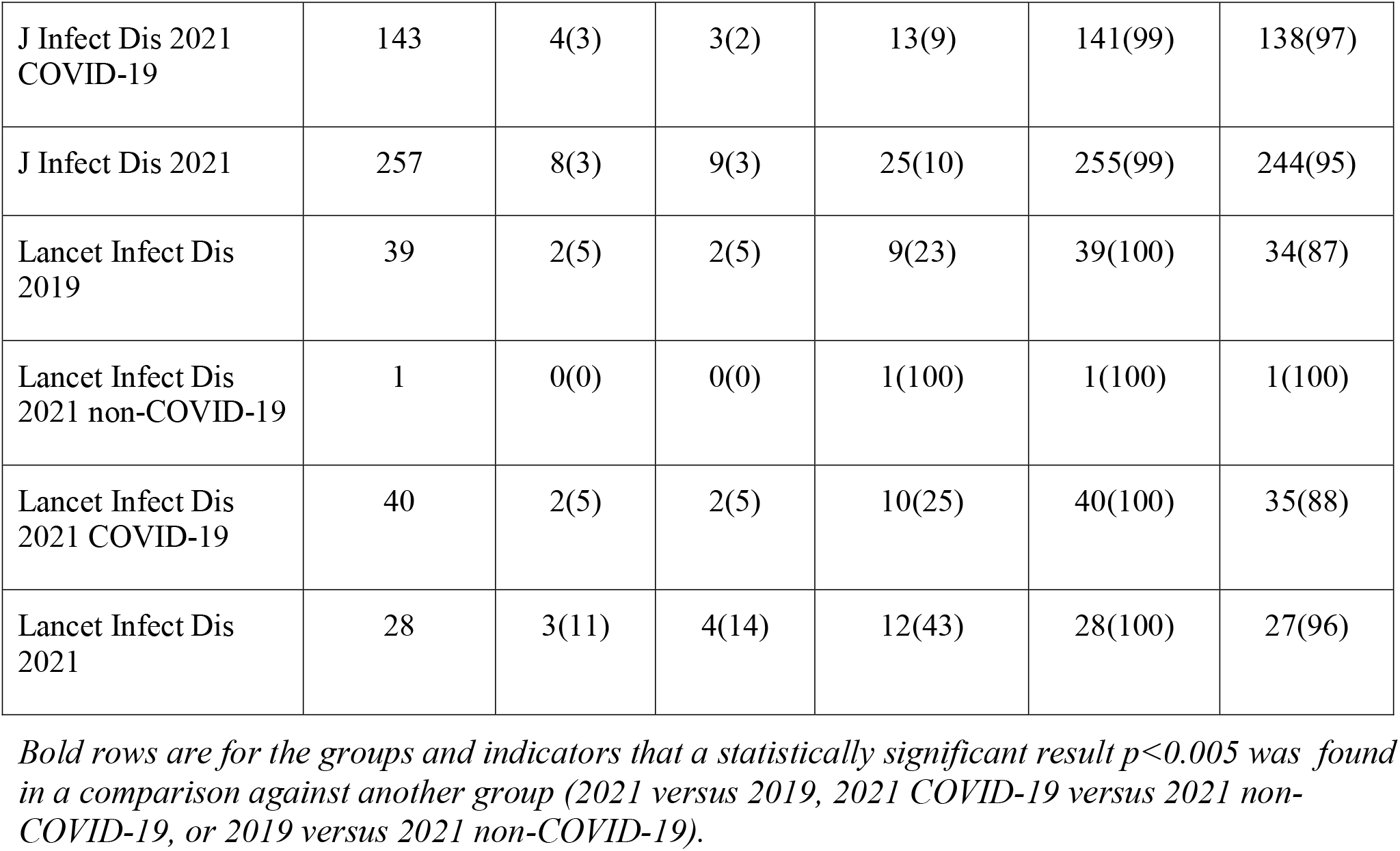
Transparency indicators stratified for Journal, Year and COVID-19 status.

| Journal, Year and COVID-19 status | Total | Code sharing<br>N(%) | Data sharing<br>N(%) | Registration<br>N(%) | Conflict of Interest<br>N(%) | Funding<br>N(%) |
| --- | --- | --- | --- | --- | --- | --- |
| BMC Infect Dis 2019 | 907 | 7(1) | 104(11) | 84(9) | 907(100) | 907(100) |
| BMC Infect Dis 2021 non-COVID-19 | 757 | 12(2) | 78(10) | 66(9) | 757(100) | 757(100) |
| BMC Infect Dis 2021 COVID-19 | 292 | 9(3) | 17(6) | 27(9) | 292(100) | 292(100) |
| BMC Infect Dis 2021 | 1049 | 21(2) | 95(9) | 93(9) | 1049(100) | 1049(100) |
| Clin Infect Dis 2019 | 233 | 0(0) | 11(5) | 0(0) | **233(100)** | **225(97)** |
| Clin Infect Dis 2021 non-COVID-19 | 248 | 5(2) | **19(8)** | **48(19)** | **248(100)** | **236(95)** |
| Clin Infect Dis 2021 COVID-19 | 369 | 11(3) | **10(3)** | **18(5)** | **343(93)** | **315(85)** |
| Clin Infect Dis 2021 | 617 | 16(3) | 29(5) | 66(11) | **591(96)** | **551(89)** |
| Clin Microbiol Infect 2019 | 22 | 0(0) | 2(9) | 3(14) | 20(91) | 20(91) |
| Clin Microbiol Infect 2021 non-COVID-19 | 11 | 0(0) | 2(18) | 1(9) | 7(64) | 10(91) |
| Clin Microbiol Infect 2021 COVID-19 | 161 | 0(0) | 7(4) | 19(12) | 116(72) | 137(85) |
| Clin Microbiol Infect 2021 | 172 | 0(0) | 9(5) | 20(12) | 123(72) | 147(85) |
| Emerg Infect Dis 2019 | 354 | 5(1) | 62(18) | 3(1) | 21(6) | 243(69) |
| Emerg Infect Dis 2021 non-COVID-19 | 306 | 1(0) | **62(20)** | 1(0) | 20(7) | 186(61) |
| Emerg Infect Dis<br>2021 COVID-19 | 252 | 8(3) | **12(5)** | 2(1) | 20(8) | 160(63) |
| Emerg Infect Dis<br>2021 | 558 | 9(2) | 74(13) | 3(1) | 40(7) | 346(62) |
| Infect Immun 2019 | 57 | 1(2) | 11(19) | 0(0) | 29(51) | 56(98) |
| Infect Immun 2021<br>non-COVID-19 | 50 | 1(2) | 16(32) | 1(2) | 28(56) | 50(100) |
| Infect Immun 2021<br>COVID-19 | 0 | 0 | 0 | 0 | 0 | 0 |
| Infect Immun 2021 | 50 | 1(2) | 16(32) | 1(2) | 28(56) | 50(100) |
| Int J Infect Dis 2019 | 35 | 0(0) | 4(11) | 1(3) | 35(100) | 35(100) |
| Int J Infect Dis 2021<br>non-COVID-19 | 37 | 0(0) | 4(11) | 3(8) | 36(97) | 37(100) |
| Int J Infect Dis 2021<br>COVID-19 | 513 | 12(2) | 19(4) | 22(4) | 498(97) | 499(97) |
| Int J Infect Dis 2021 | 550 | 12(2) | 23(4) | 25(5) | 534(97) | 536(97) |
| J Antimicrob<br>Chemother 2019 | 86 | 1(1) | 14(16) | 9(10) | 28(33) | 84(98) |
| J Antimicrob<br>Chemother 2021<br>non-COVID-19 | 101 | 1(1) | 13(13) | 8(8) | 42(42) | 98(97) |
| J Antimicrob<br>Chemother 2021<br>COVID-19 | 25 | 0(0) | 2(8) | 3(12) | 7(28) | 23(92) |
| J Antimicrob<br>Chemother 2021 | 126 | 1(1) | 15(12) | 11(9) | 49(39) | 121(96) |
| J Infect Dis 2019 | 125 | 3(2) | 9(7) | 21(17) | 125(100) | **125(100)** |
| J Infect Dis 2021<br>non-COVID-19 | 114 | 4(4) | 6(5) | 12(11) | 114(100) | **106(93)** |
| J Infect Dis 2021<br>COVID-19 | 143 | 4(3) | 3(2) | 13(9) | 141(99) | 138(97) |
| J Infect Dis 2021 | 257 | 8(3) | 9(3) | 25(10) | 255(99) | 244(95) |
| Lancet Infect Dis<br>2019 | 39 | 2(5) | 2(5) | 9(23) | 39(100) | 34(87) |
| Lancet Infect Dis<br>2021 non-COVID-19 | 1 | 0(0) | 0(0) | 1(100) | 1(100) | 1(100) |
| Lancet Infect Dis<br>2021 COVID-19 | 40 | 2(5) | 2(5) | 10(25) | 40(100) | 35(88) |
| Lancet Infect Dis<br>2021 | 28 | 3(11) | 4(14) | 12(43) | 28(100) | 27(96) |

*Bold rows are for the groups and indicators that a statistically significant result $p < 0.005$ was found* *in a comparison against another group (2021 versus 2019, 2021 COVID-19 versus 2021 non-* *COVID-19, or 2019 versus 2021 non-COVID-19).*

### 3.3 Manually assessed sample of 200 randomly selected articles

In the random sample of 200 articles that were examined in depth, the most common countries of first and last author were US and China and the collectively accounted for about a third of the articles. Most studies were classified as observational (34%) or epidemiologic surveillance (22%). The large majority of studies (88%) addressed a specific pathogen. Most studies (55%) used neither p-values nor confidence intervals in their abstract. Most studies were on humans (80%) and age groups represented were very diverse. 166 of the 200 (83%) studies could be characterized as clinical studies Among the clinical studies, 42 (25%) were interventional and 124 (75%) were non-interventional. The primary focus of the study was epidemiology in 76 (38%) articles, therapy in 34 (17%), diagnosis in 21 (11%), and prevention in 14 (7%) articles. 13 (7%) had a focus on risk and 9 (5%) had a pathophysiological focus. Details on the characteristics of the manually assessed random sample appear in the Supplementary Table 2.

The same supplementary table 2 also shows details on the presence of code sharing, data sharing, and registration in the 200 papers, according to these characteristics. As shown, no characteristic seemed to be associated with markedly higher rates of code sharing or data sharing, perhaps with the exception of code sharing for predictive models (28%), but numbers are very thin to make robust inferences. For registration, 10/10 clinical trials (100%) and 4/13 (31%) of systematic reviews were registered, but only 3 other studies were registered among the remaining 177 articles.

### 3.4 Manual validation

The rate of false positives of the automated algorithms was 0/3 (0%) for code sharing, 1/12 (8%) for data sharing, 1/19 (5%) for registration, 0/145 (0%) for conflict of interest disclosures and 0/166 (0%) for funding disclosures On the other hand the false negative rate was 2/180 (1%) for code sharing, 4/171 (2%) for data sharing, 1/164 (1%) for registration, 1/38 (3%) for conflict of interest disclosures and 1/17 (6%) for funding disclosures. 41/182 (23%) of funding disclosures practically stated that there was no funding for the study. 117/162 (72%) of the conflict of interest statements practically declared that there was no conflict of interest.

Adjusting for the manual validation results, the corrected proportions of transparency indicators were 3% for code sharing, 11% for data sharing, 8% for registrations, 79% for conflict of interest disclosures and 92% for funding disclosures.

## 4 Discussion

This evaluation of 5340 articles in infectious disease research published in 2019 and in 2021 in the top 9 specialty journals with respects to impact showed that only a small minority of articles shared code or data. Study registration was also rare, and it pertained almost exclusively to clinical trials and some systematic reviews. Conflict of interest and funding disclosures were, conversely, very common. There was no major change in absolute magnitude in the proportion of articles that satisfied these transparency indicators during 2021 (a pandemic year) versus 2019 (a pre-pandemic years). COVID-19 articles had modestly lower rates of data sharing.

In a previous evaluation(11) we had studied 1338 articles from 2019 and 2021 and we had showed that approximately a quarter of publications shared code, and a modestly higher proportion shared data in studies of infectious disease modelling. High rates of conflict of interest and funding disclosures were also observed in the same study (around 90% for both). Registrations on the other hand were below 1% in that sample. The disparities are likely due to the difference in the study characteristics; infectious disease models were very sparse in our sample. Most of the studies that we evaluated were clinical, and since registration is more common in clinical trials and clinical research overall(21) this has likely affected the overall average in our included sample. However, registration is distinctly uncommon outside of clinical trials, with the exception of some systematic reviews.

Moreover, code and data sharing are expected to be more common in modeling studies, where such sharing is indispensable, while these research practices remain quite rare in the bulk of clinical and epidemiological research that comprises the vast volume of papers published in infectious disease journals.

The numbers regarding code sharing are similar to the overall assessments previously performed by Serghiou et al.(17), but the rate of data sharing and registration was higher when studying the entire PubMed Central Open Access subset across all scientific disciplines. Iqbal et al.(22), showed similar rates of transparency in publications indexed in 2014 when they studied a random sample of 441 PubMed articles between 2000 and 2014. Overall, infectious disease specialty journals seem to be performing below the average of biomedical journals in data sharing and registration.

Code and data sharing are critical elements of computational reproducibility. They are an essential part of most conducted research as it allows the reanalysis and the assessment of the methods section for potential errors or non-disclosed analytical approaches that may affect the study results. We should acknowledge that code sharing may not be applicable to some types of infectious disease articles where there are no quantitative computational parts. Also data sharing may need to take into account the specific circumstances of the research, e.g. consent requirements and the need for deidentification, that may be common in infectious disease research, especially clinical studies.

Nevertheless, more transparent sharing in general is desirable. Sharing practices can also maximize the future use of the data and enhance the value of the collected information(23). The absence of sharing affects the trustworthiness of the analytical approaches in published research. Many efforts have been directed toward increasing reproducibility through increased sharing. These include the changes in journal policy for Science regarding data sharing and the new regulations regarding data sharing that the NIH aims to implement in 2023 (8,24,25), further showing the importance of this aspect of transparency.

Registration, on the other hand, is key in research that aims to inform clinical practice and policy and to avoid “vibration of effects”(26,27), i.e. instability in the results due to post-hoc, selective choices of statistical analyses and reporting bias, choosing which analyses and outcomes to report. A sensible prospective registration process may increase the reliability of the results from observational as well as interventional research and one may compare notes between registered protocols and subsequent results that are made available(10,28–31). Nevertheless, registration remains very uncommon outside of randomized trials and systematic reviews.

### 4.1 Limitations

Our meta-research assessment has certain limitations. First, the existence of transparency indicators in the text of a published article does not guarantee the informational value of the statements. For example, for code sharing and data sharing, one would have to examine in depth whether the code and the data are not only accessible but functional and contain all the information needed to use them.

Second, the veracity of some statements of transparency may also be brought into question, especially for conflict of interest and funding disclosures. Disclosures are notoriously difficult to verify for their completeness and accuracy, despite the emerging availability of some resources such as the Centers for Medicare & Medicaid Services Open Payments database(32) in the US. Many assessments have been performed with this data and shown a worrying amount of undisclosed conflicts in opioid prescription(33), dermatology(12) and otolaryngology(13) guidelines among others. The research and clinical practice of infectious diseases is not exempt from this pattern. One well known example is the so called “Lyme wars”, where conflicts remained unreported and affected developed clinical practice guidelines(34); this also led to a class action lawsuit and an investigation by the Attorney General that forced IDSA to redo the Lyme guidelines. Other empirical evaluations have also shown large rates of conflicts in infectious diseases in specific settings or countries, e.g. in Japan (35).

Second, infectious disease journals vary a lot in scope and types of studies that they publish. Infectious disease research spans a wide range of investigations, from in vitro studies of mice to clinical research. Transparency and reproducibility challenges may differ for different types of research. We used a large sample and diverse journals so as to capture this diversity, but some types of studies may have been under-represented. The 9 examined journals also publish a lion’s share of the papers published by infectious disease specialty journals (as characterized by JCR). However, some very influential infectious disease-related studies are not published in the specialty journals of the field, but in general medical and general science journals (e.g. Nature or Science) and many papers on infections may appear in specialty journals outside of those focusing on infections. Transparency indicators may exhibit different patterns in these journals.

Third, only PMC OA subset publications were included and not the full list of PubMed publications from the 9 journals. There may be specific tendencies in the open access subset that leads to a misrepresentation of proportions of transparency. Open access articles may be more transparent overall; if so, we may have overestimated the rate of transparency in the field. Furthermore, the overall rate of open access status has increased with the COVID-19 pandemic and the funders’ and publishers’ open sharing commitments that came as a response to the pandemic. Prior estimates have shown upward of 97% of COVID-19 papers being open access publications(36–38), outpacing any other field’s openness and further skewing the sample.

Fourth, we sampled from two recent years, 2019 and 2021, where 2021 is a unique pandemic year and therefore these numbers are possible to not reflect true yearly trends but mostly the covidization of science that has been observed across fields and disciplines and its repercussions(5,7,20,39). However, we did use a comparison focusing only on the non-COVID-19 papers in these two years.

Finally, some infectious disease studies are purely non-quantitative and coding analyses would make little or no sense. Also registration is not necessarily feasible or appropriate for all studies. Therefore, one should not expect these transparency indicators to be feasible to attain in 100% of the published infectious disease articles, even under perfect transparency settings.

### 4.2 Conclusions

The repeated disconcerting finding of opacity in medical research and the reproducibility crisis may be inextricably joined. Lack of transparency poses obstacles to reliable and rigorous scientific research. Therefore, infectious disease specialty journals would benefit from more common adoption of code and data sharing for the articles that they publish. Registration practices could also be optimized in the field.

## Supporting information

Manually extracted characteristics

## Data Availability

The code and data generated and analyzed for this study is available in https://doi.org/10.17605/OSF.IO/FYZPX (for the code and datasets, as well as the protocol).

## 7 Acknowledgments

None

