## Supplemental figures and tables for "Transparency in infectious disease research: a meta-research survey of specialty journals"

Supplementary Material

### 1 Supplementary Figures and Tables

#### 1.1 Supplementary Figures

#### Supplementary Figure 1. Forest plot: Fixed effects meta-analysis of code sharing across journal, comparing papers from 2021 to 2019

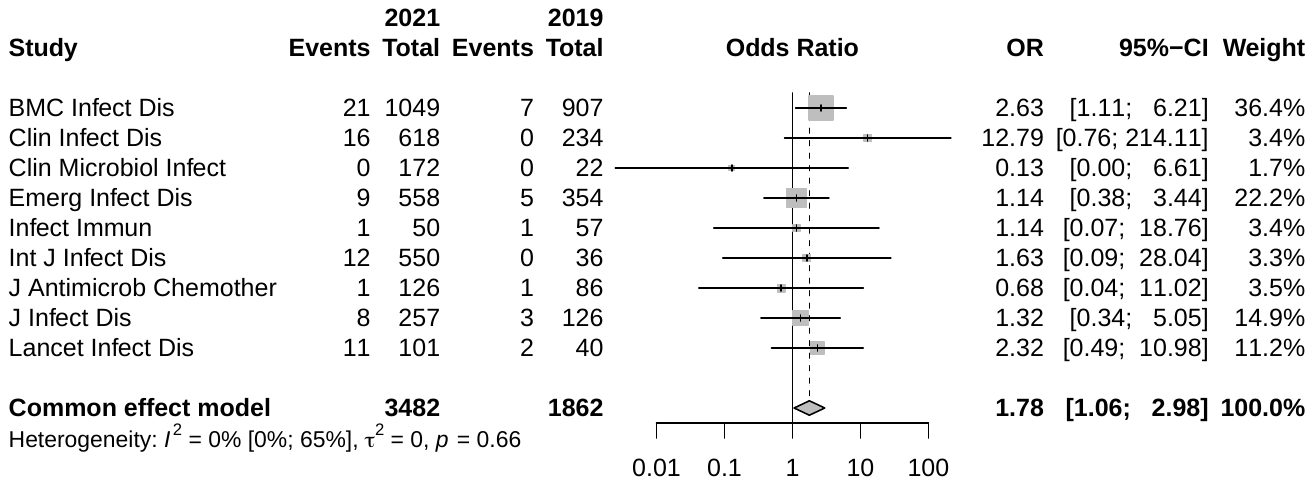

#### Supplementary Figure 2. Forest plot: Fixed effects meta-analysis of code sharing across journals, comparing non-COVID-19 papers from 2021 to ones from 2019

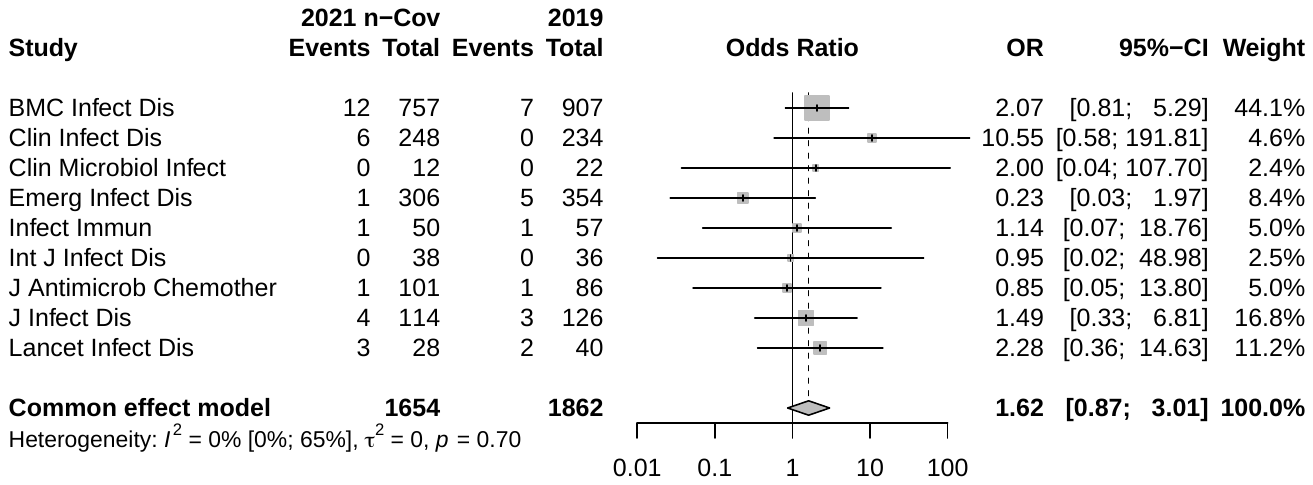

#### Supplementary Figure 3. Forest plot: Fixed effects meta-analysis of code sharing across journal, comparing COVID-19 to non-COVID-19 papers from 2021

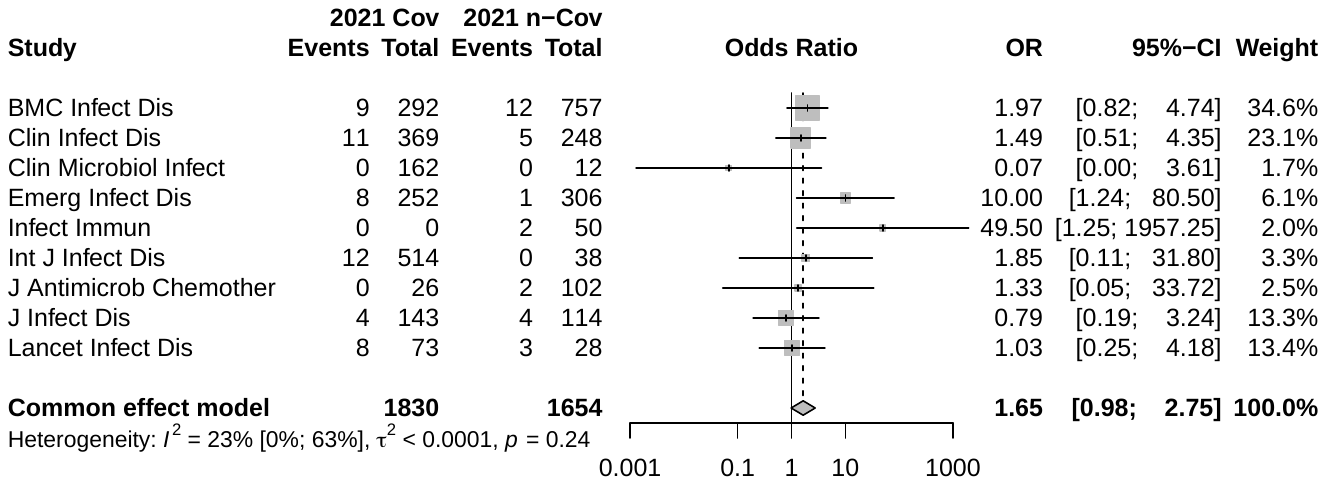

**Supplementary Figure 4.** Forest plot: Fixed effects meta-analysis of data sharing across journal, comparing papers from 2021 to 2019

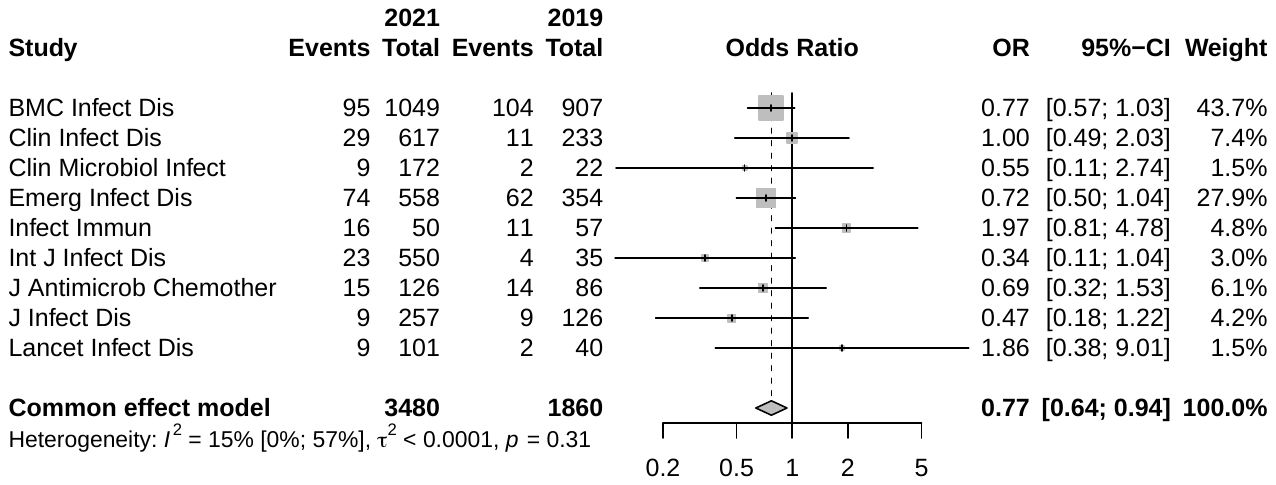

**Supplementary Figure 5.** Forest plot: Fixed effects meta-analysis of data sharing across journal, comparing non-COVID-19 papers from 2021 to ones from 2019
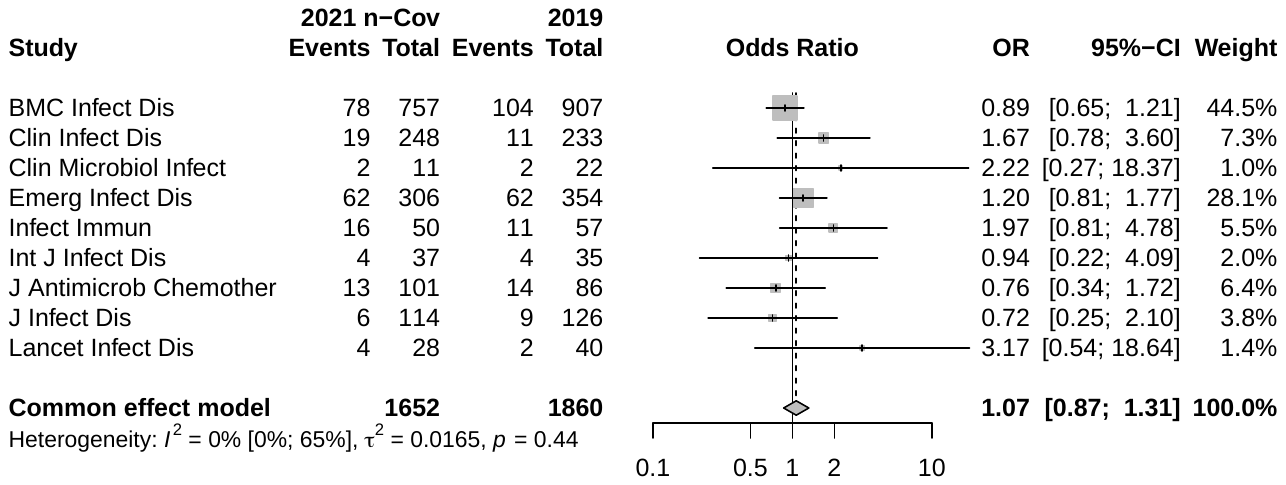

**Supplementary Figure 6.** Forest plot: Fixed effects meta-analysis of data sharing across journal, comparing COVID-19 to non-COVID-19 papers from 2021
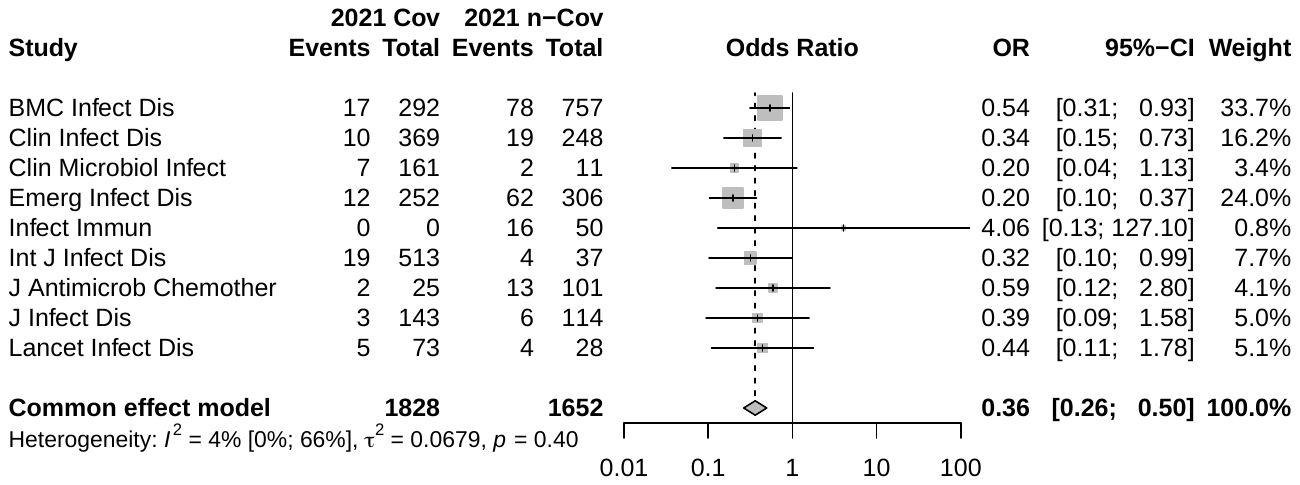

**Supplementary Figure 7.** Forest plot: Fixed effects meta-analysis of registrations across journal, comparing papers from 2021 to 2019

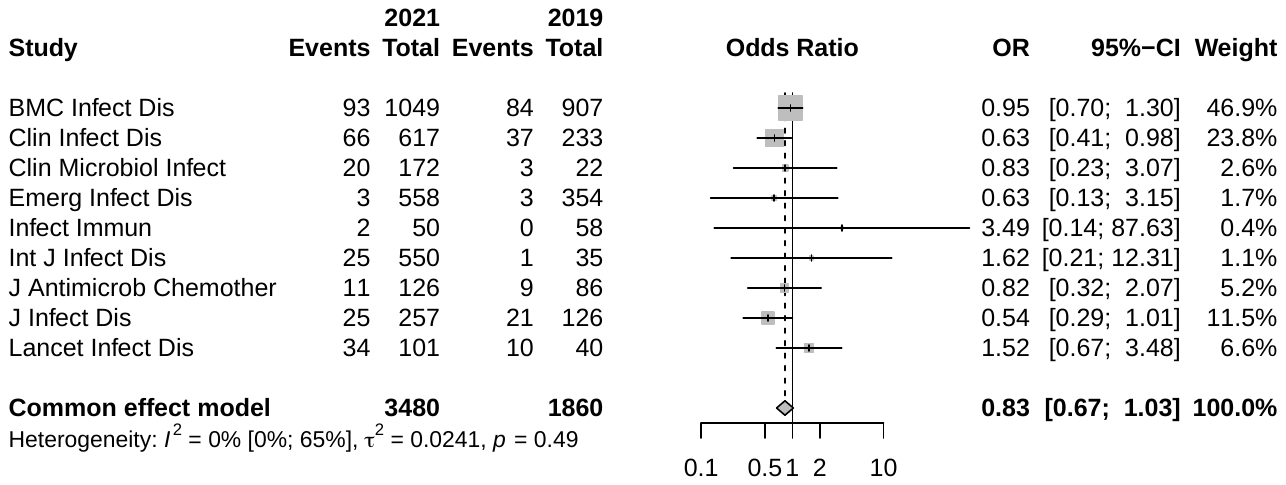

**Supplementary Figure 8.** Forest plot: Fixed effects meta-analysis of registrations across journal, comparing non-COVID-19 papers from 2021 to ones from 2019

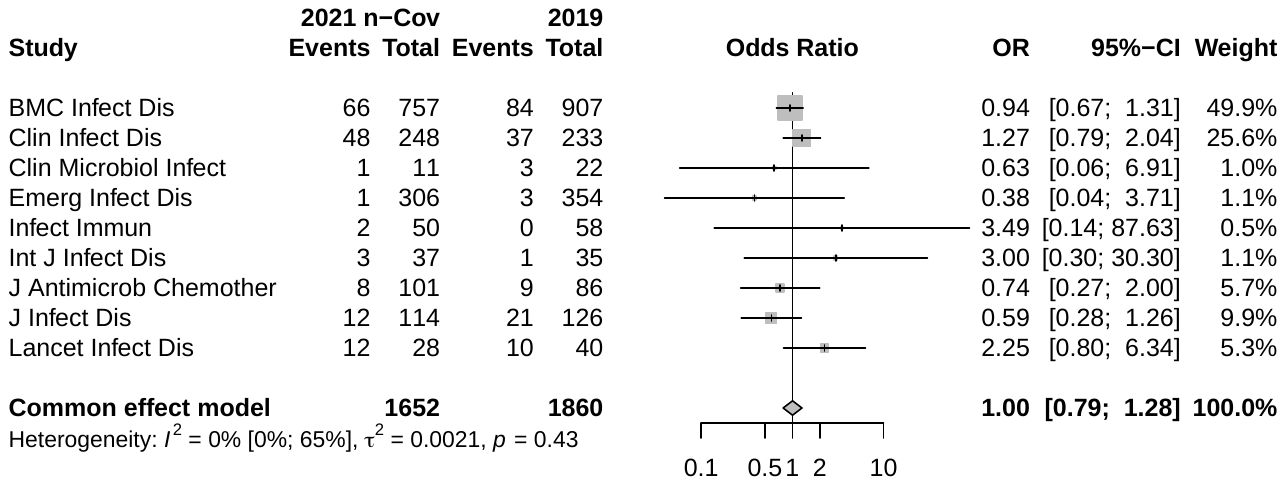

**Supplementary Figure 9.** Forest plot: Fixed effects meta-analysis of registrations across journal, comparing COVID-19 to non-COVID-19 papers from 2021

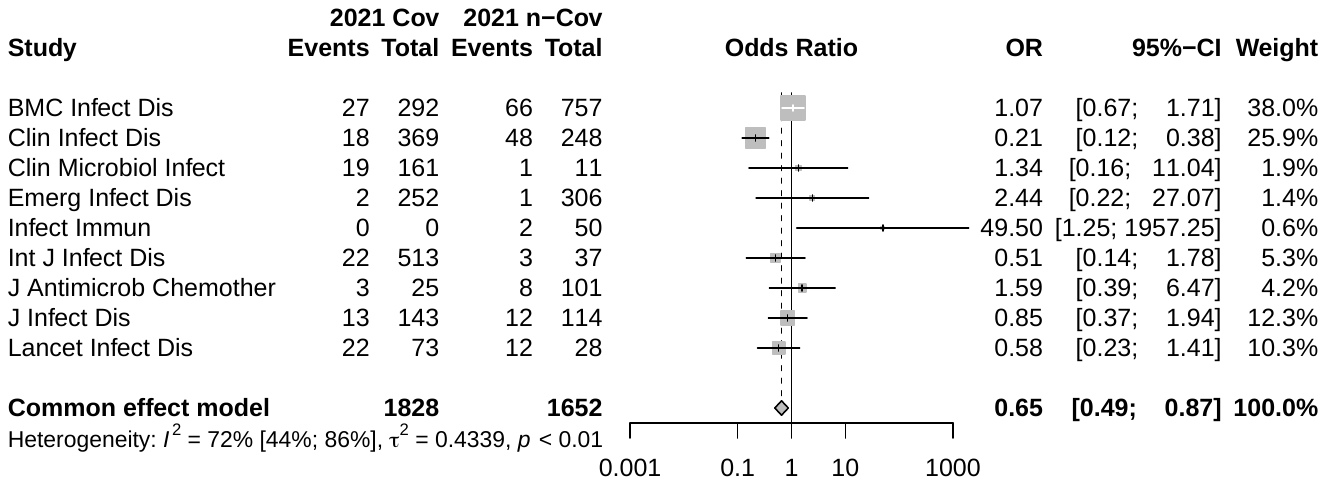

**Supplementary Figure 10.** Forest plot: Fixed effects meta-analysis of conflict of interest statements across journal, comparing papers from 2021 to 2019
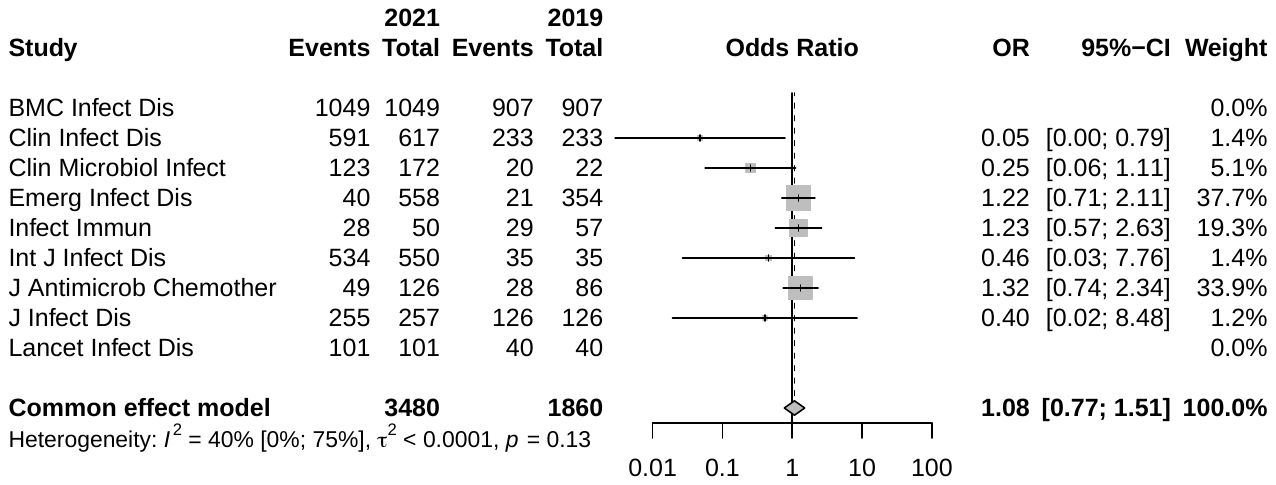

**Supplementary Figure 11.** Forest plot: Fixed effects meta-analysis of conflict of interest statements across journal, comparing non-COVID-19 papers from 2021 to ones from 2019
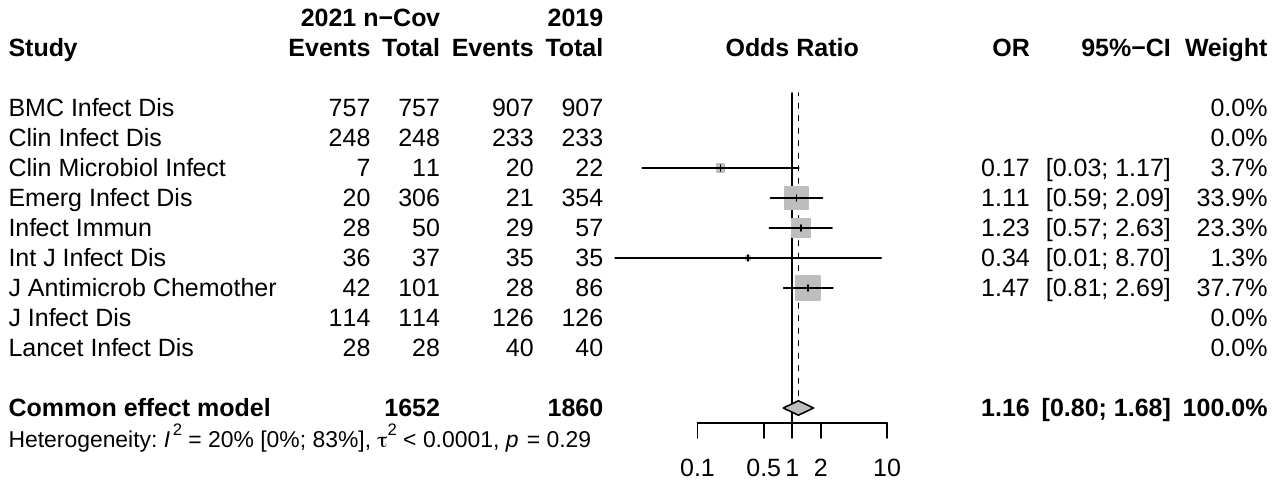

**Supplementary Figure 12.** Forest plot: Fixed effects meta-analysis of conflict of interest statements across journal, comparing COVID-19 to non-COVID-19 papers from 2021
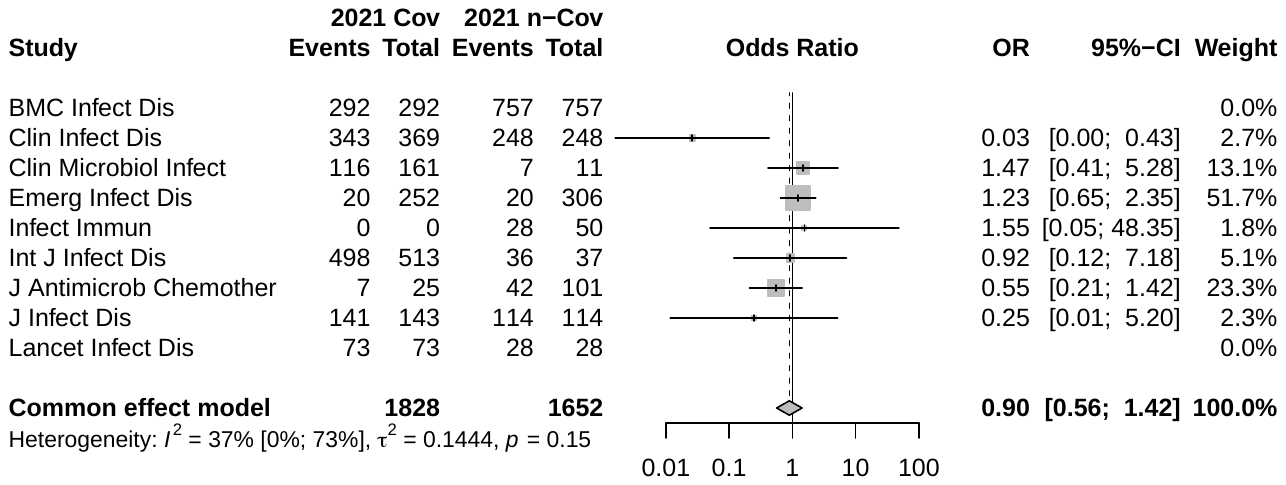

**Supplementary Figure 13.** Forest plot: Fixed effects meta-analysis of funding statement across journal, comparing papers from 2021 to 2019
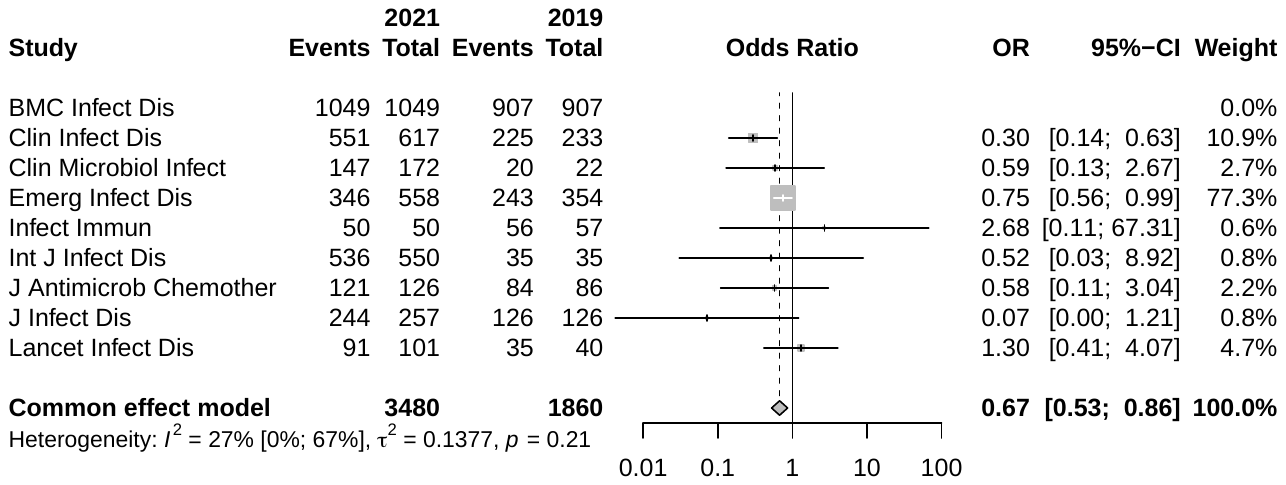

**Supplementary Figure 14.** Forest plot: Fixed effects meta-analysis of funding statement across journal, comparing non-COVID-19 papers from 2021 to ones from 2019
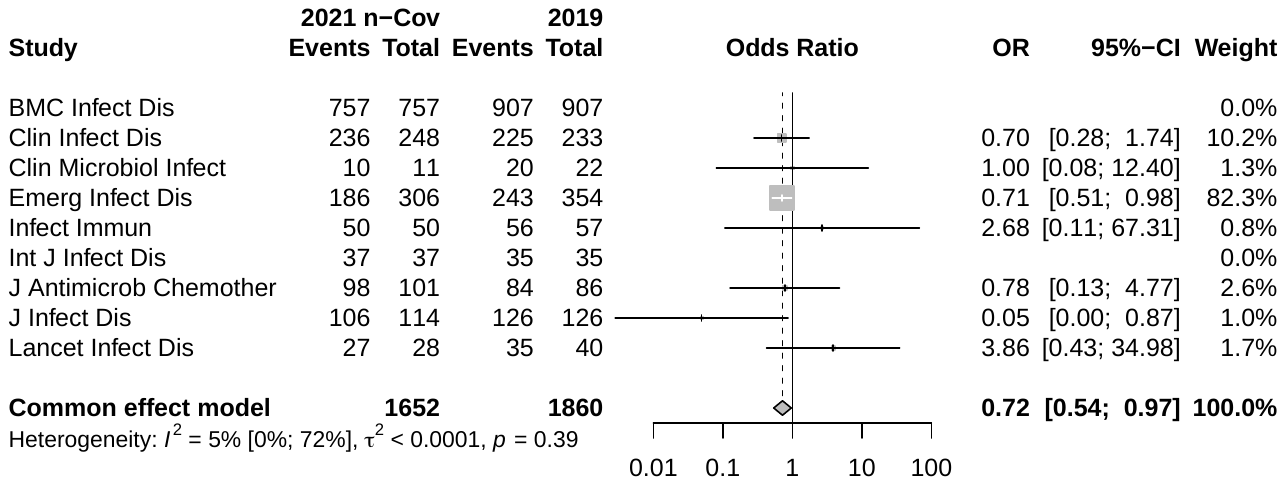

**Supplementary Figure 15.** Forest plot: Fixed effects meta-analysis of funding statement across journal, comparing COVID-19 to non-COVID-19 papers from 2021
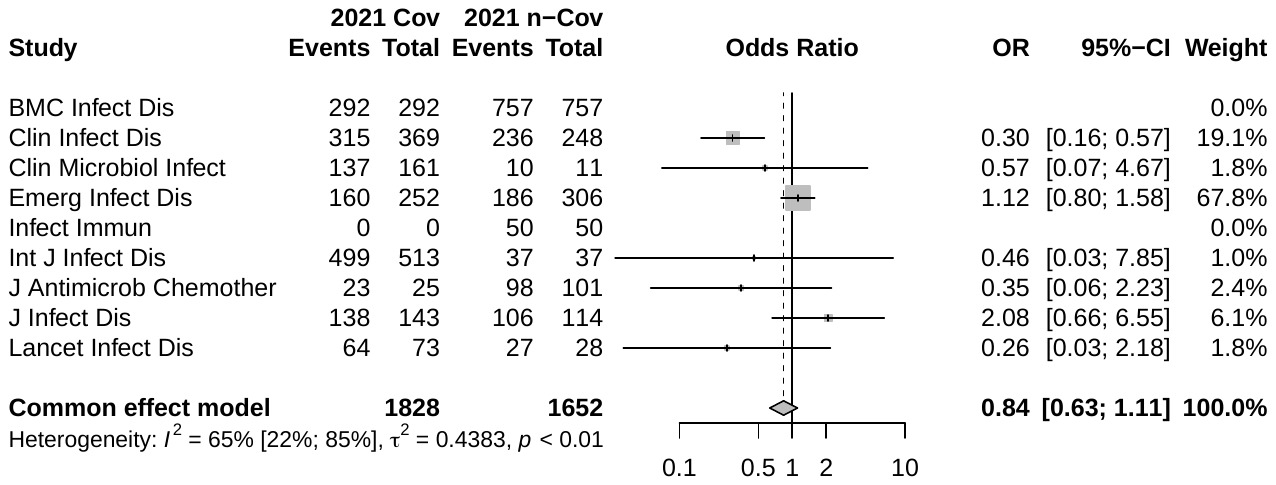

#### Supplementary Figure 16. Forest plot: Random effects meta-analysis of code sharing across journal, comparing papers from 2021 to 2019

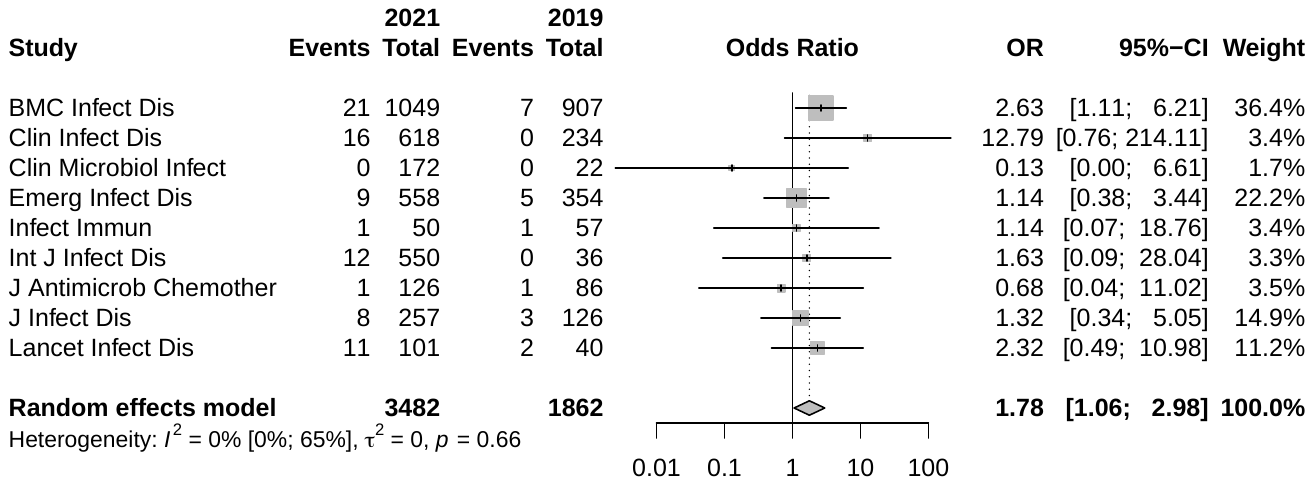

#### Supplementary Figure 17. Forest plot: Random effects meta-analysis of code sharing across journal, comparing non-COVID-19 papers from 2021 to ones from 2019

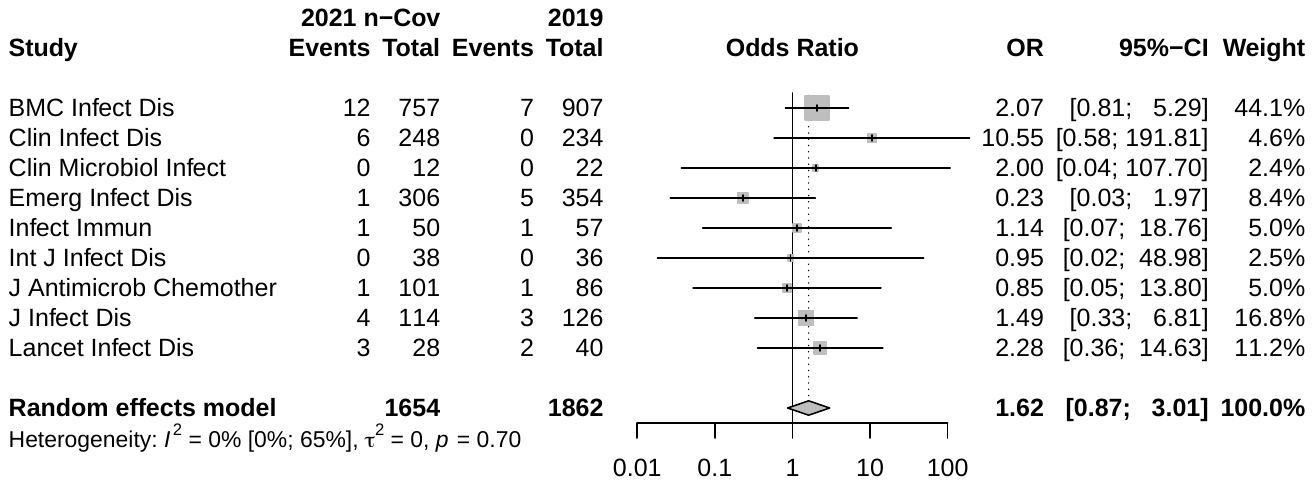

**Supplementary Figure 18.** Forest plot: Random effects meta-analysis of code sharing across journal, comparing COVID-19 to non-COVID-19 papers from 2021
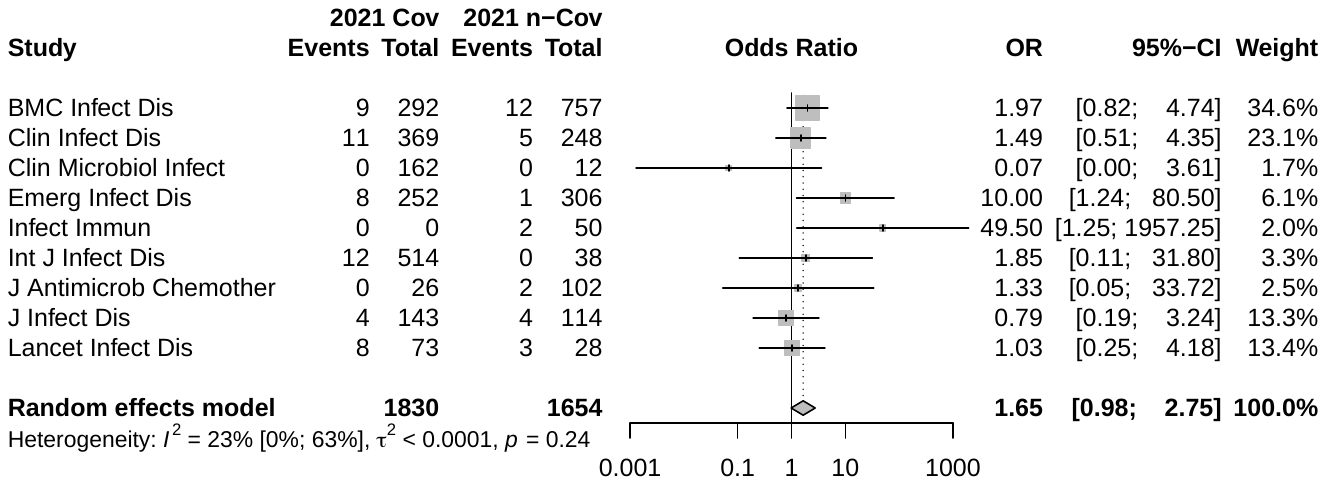

**Supplementary Figure 19.** Forest plot: Random effects meta-analysis of data sharing across journal, comparing papers from 2021 to 2019
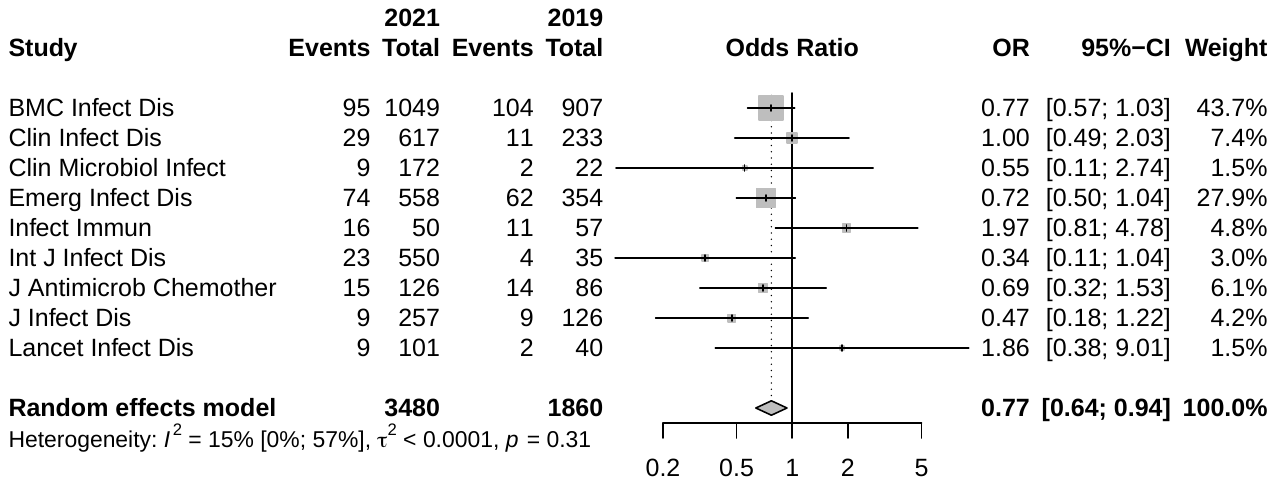

**Supplementary Figure 20.** Forest plot: Random effects meta-analysis of data sharing across journal, comparing non-COVID-19 papers from 2021 to ones from 2019
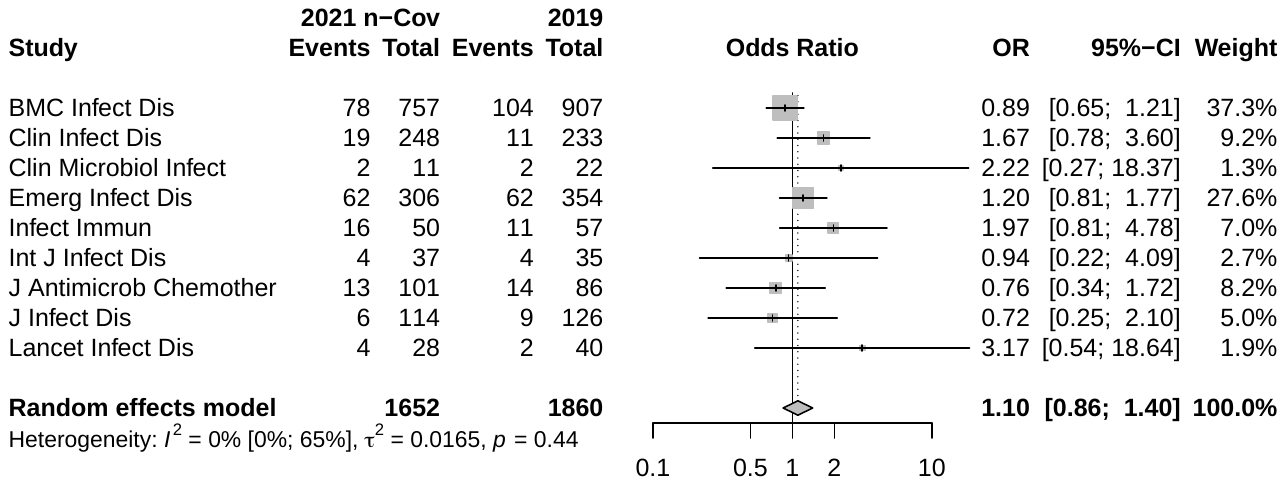

**Supplementary Figure 21.** Forest plot: Random effects meta-analysis of data sharing across journal, comparing COVID-19 to non-COVID-19 papers from 2021
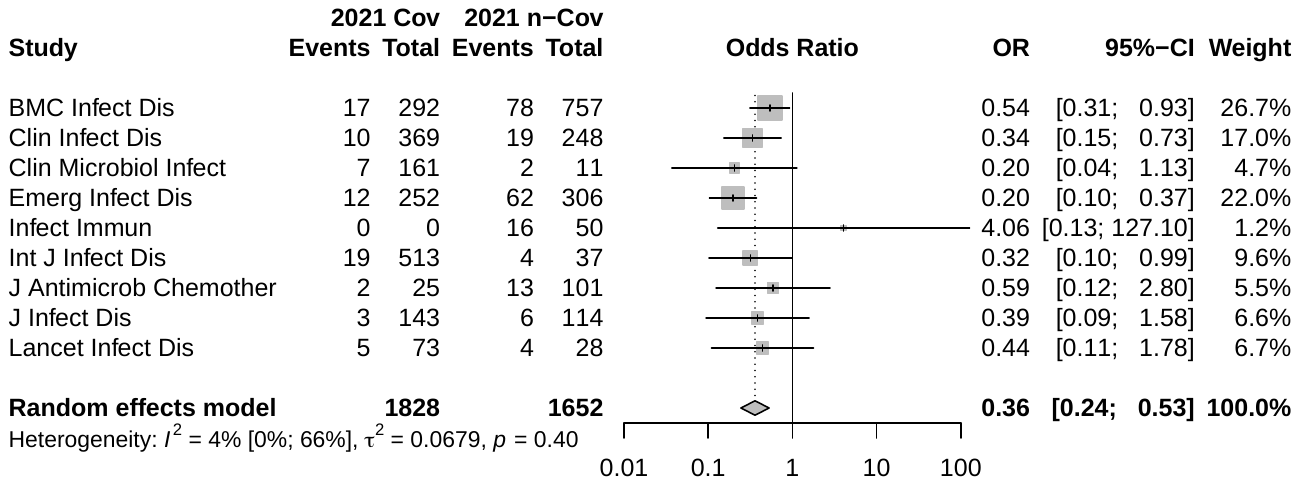

**Supplementary Figure 22.** Forest plot: Random effects meta-analysis of registrations across journal, comparing papers from 2021 to 2019
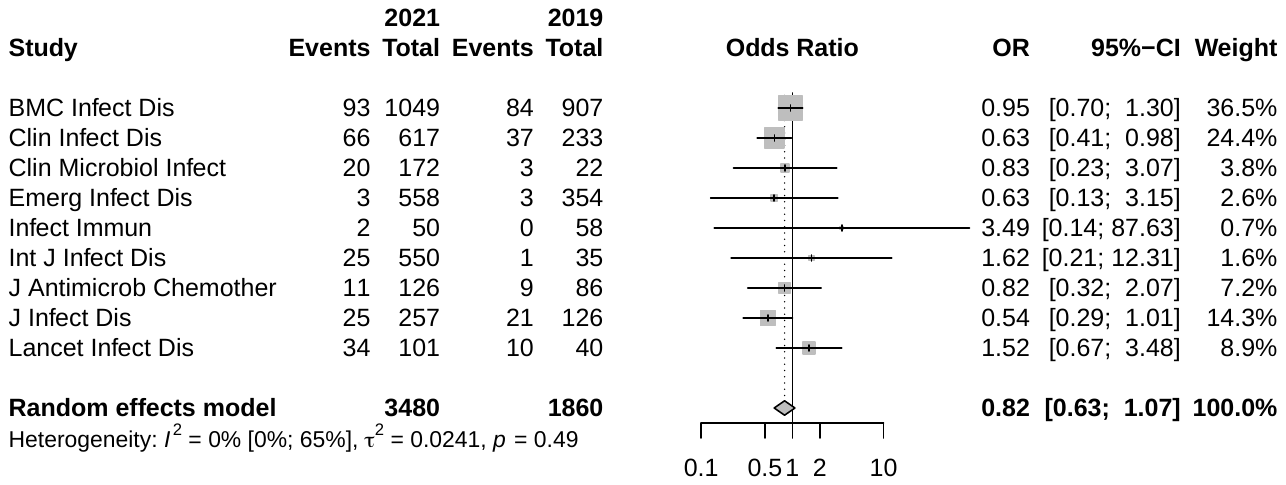

**Supplementary Figure 23.** Forest plot: Random effects meta-analysis of registrations across journal, comparing non-COVID-19 papers from 2021 to ones from 2019
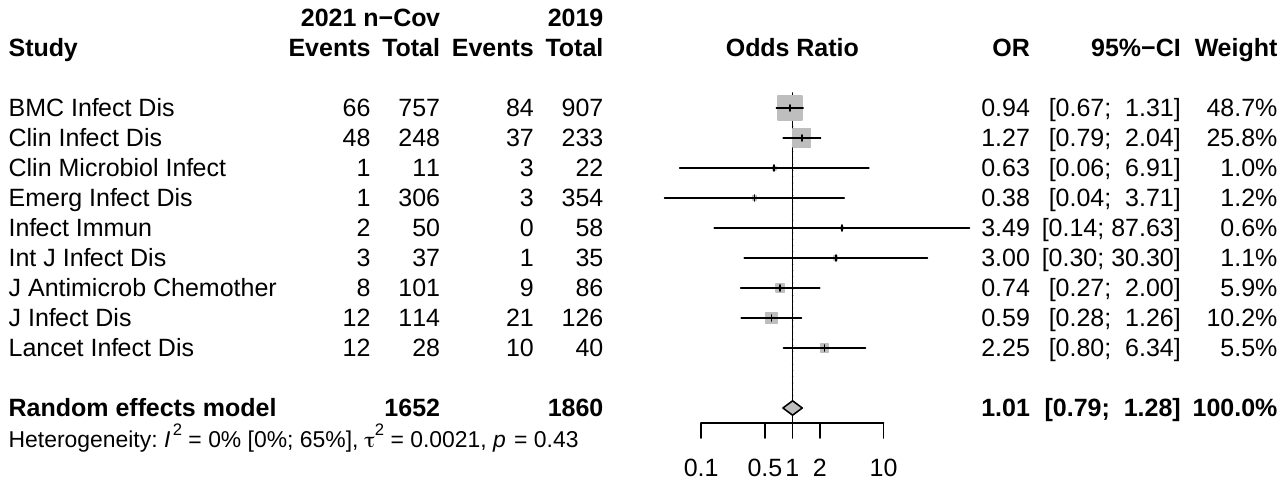

**Supplementary Figure 24.** Forest plot: Random effects meta-analysis of registrations across journal, comparing COVID-19 to non-COVID-19 papers from 2021
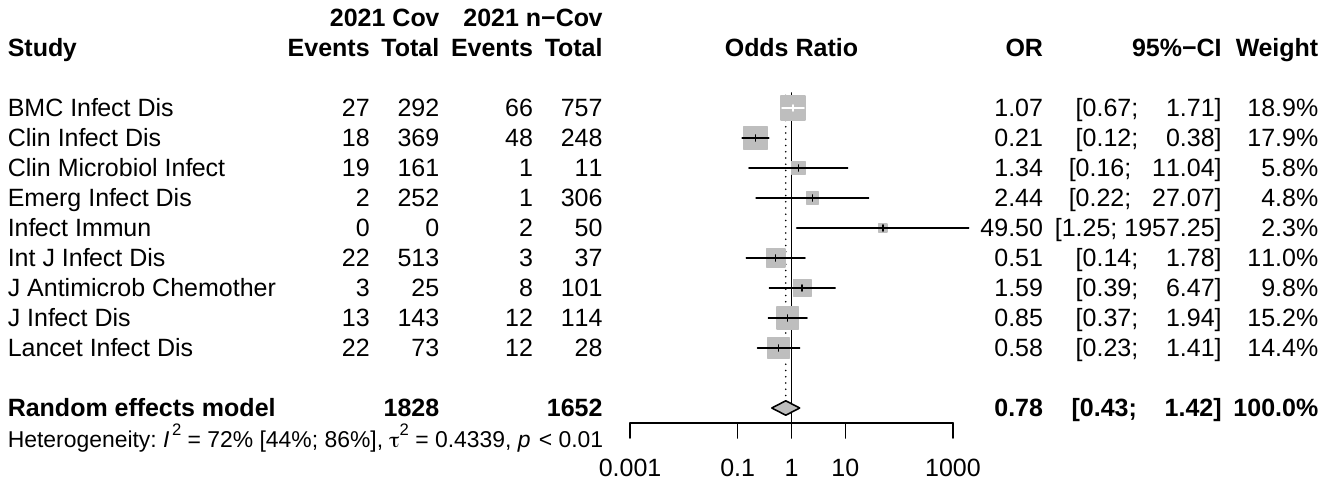

**Supplementary Figure 25.** Forest plot: Random effects meta-analysis of conflict of interest statements across journal, comparing papers from 2021 to 2019
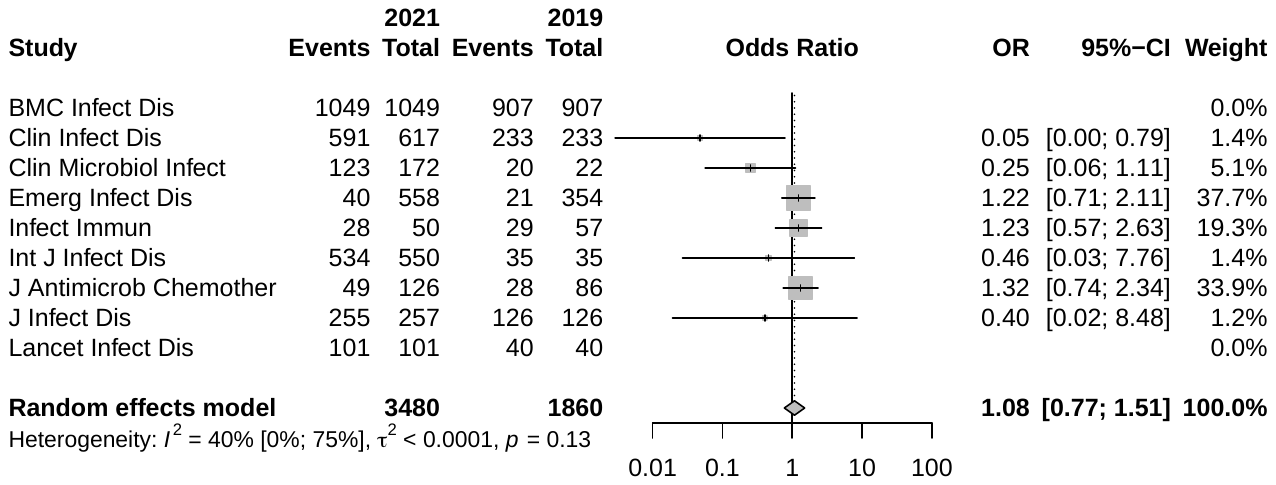

**Supplementary Figure 26.** Forest plot: Random effects meta-analysis of conflict of interest statements across journal, comparing non-COVID-19 papers from 2021 to ones from 2019
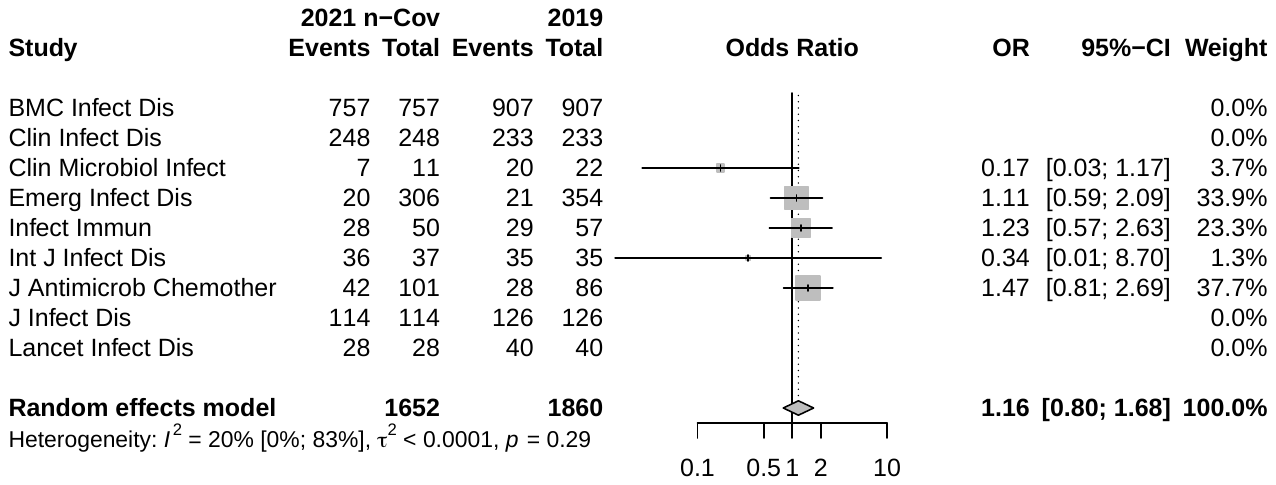

**Supplementary Figure 27.** Forest plot: Random effects meta-analysis of conflict of interest statements across journal, comparing COVID-19 to non-COVID-19 papers from 2021
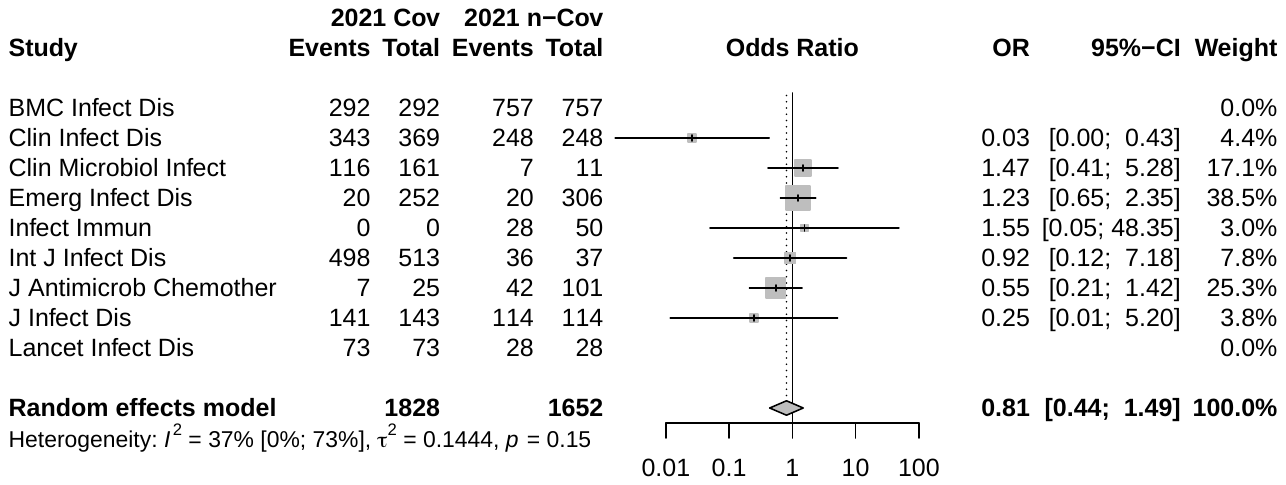

**Supplementary Figure 28.** Forest plot: Random effects meta-analysis of funding statement across journal, comparing papers from 2021 to 2019
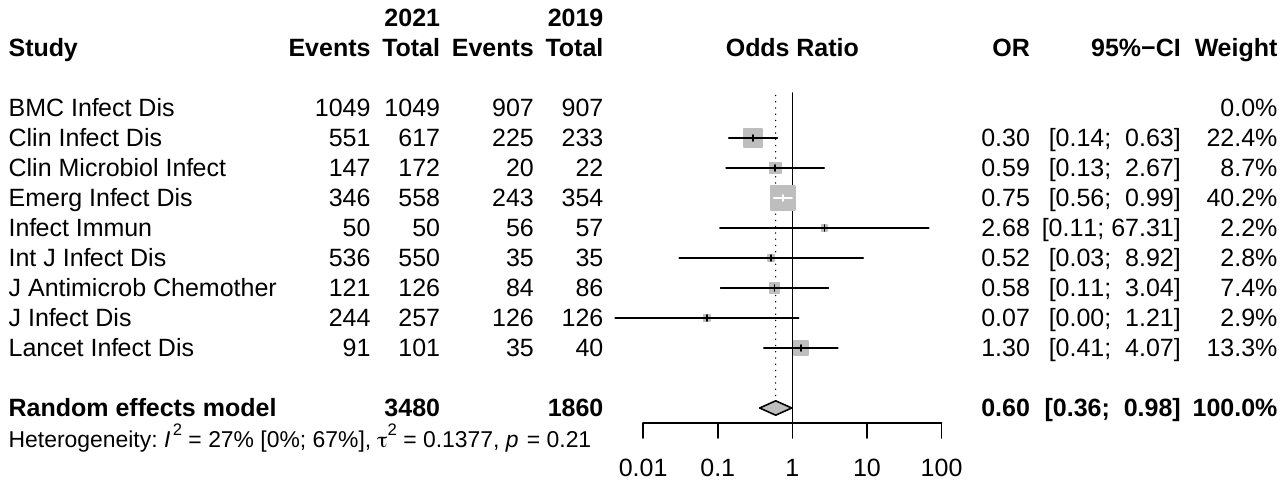

**Supplementary Figure 29.** Forest plot: Random effects meta-analysis of funding statement across journal, comparing non-COVID-19 papers from 2021 to ones from 2019
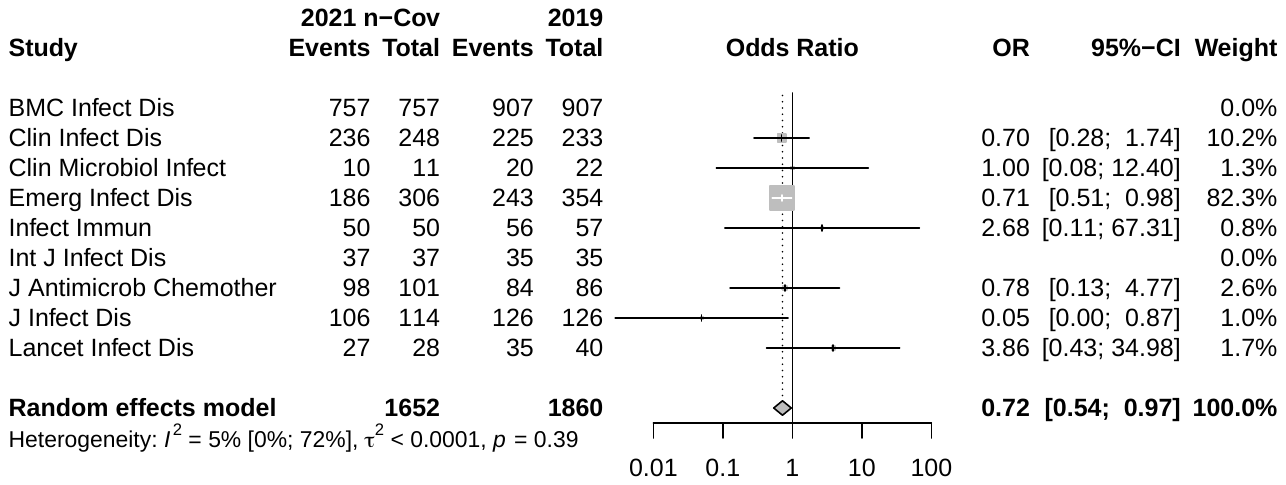

**Supplementary Figure 30.** Forest plot: Random effects meta-analysis of funding statement across journal, comparing COVID-19 to non-COVID-19 papers from 2021
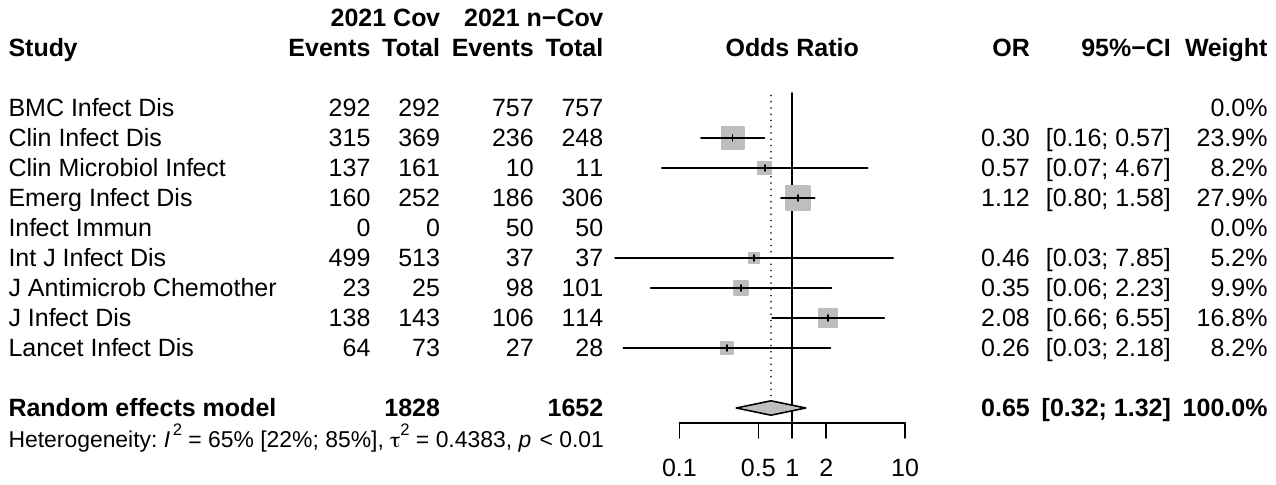

#### 1.2 Supplementary Tables

**Supplementary Table 1.** 9 Journals with most citations

| Journal name | Number of citations |
| --- | --- |
| Clinical Infectious Diseases | 115,949 |
| Journal of Infectious Diseases | 58,677 |
| Lancet Infectious Diseases | 52,835 |
| Infection and Immunity | 51,122 |
| Emerging Infectious Diseases | 48,544 |
| Journal of Antimicrobial Chemotherapy | 40,176 |
| Clinical Microbiology and Infection | 30,884 |
| BMC Infectious Diseases | 28,296 |
| International Journal of Infectious Diseases | 26,024 |

**Supplementary Table 2.** Article characteristics and transparency indicators (based on random sample of 200 articles)

| **Meta-data and statistical presentation in abstract** | **Total of subgroup** N(%) alt. mean (95% CI) | **Code sharing** N(%) alt. mean (95% CI) | **Data sharing** N(%) alt. mean (95% CI) | **Registration** N(%) alt. mean (95% CI) |
| --- | --- | --- | --- | --- |
| Country of first author  USA | 41(21) | 0(0) | 3(7) | 4(10) |
| China | 22(11) | 0(0) | 3(14) | 3(14) |
| Rest | 137(69) | 3(2) | 5(4) | 12(9) |
| Country of last author  USA | 46(23) | 0(0) | 1(2) | 3(7) |
| China | 22(11) | 0(0) | 3(14) | 3(14) |
| Rest | 132(66) | 3(2) | 5(2) | 12(9) |
| No. of pages | 8.28(7.8,8.8) | 9.3(1.7,16.9) | 8.25(6.0,10.5) | 9.89(7.8,11.9) |
| No. of Figures | 2.35(2.1,2.6) | 3.66(2.2,5.1) | 3.5(2.2,4.8) | 2.32(1.4,3.2) |
| No. of Tables | 2.21(2.0,2.0) | 1.66(-2.1,5.4) | 1.66(0.8,2.6) | 2.89(2.1,3.7) |
| No. of Appendices | 0.8(0.6,1.0) | 1.66(-1.2,4.5) | 1.58(-0.2,3.4) | 0.74(0.4,1.1) |
| Statistical presentation of abstract  P-values | 39(20) | 0(0) | 1(3) | 2(5) |
| Confidence interval | 58(29) | 1(2) | 1(2) | 10(17) |
| Both | 18(9) | 0(0) | 1(6) | 1(6) |
| Neither | 110(55) | 2(2) | 9(8) | 6(5) |
| **Study design** N(%) | **Total of subgroup** N(%) | **Code sharing** N(%) | **Data sharing** N(%) | **Registration** N(%) |
| Observational | 68(34) | 1(1) | 3(4) | 3(4) |
| Epidemiologic Surveillance | 43(22) | 0(0) | 2(5) | 0(0) |
| In vitro study | 19(8) | 0(0) | 3(16) | 0(0) |
| Systematic review | 13(8) | 0(0) | 2(15) | 4(31) |
| Clinical trial | 10(5) | 0(0) | 0(0) | 10(100) |
| Case reports/Case series | 10(5) | 0(0) | 1(10) | 0(0) |
| Prediction model | 7(4) | 2(30) | 0(0) | 0(0) |
| **Clinical studies** N(%) | **Total of subgroup** N(%) | **Code sharing** N(%) | **Data sharing** N(%) | **Registration** N(%) |
| Non-clinical studies | 34(17) | 0(0) | 2(6) | 0(0) |
| Clinical studies | 166(83) | 3(2) | 10(6) | 19(11) |
| Interventional | 42(25) | 0(0) | 2(5) | 14(33) |
| Non-interventional | 124(75) | 3(2) | 8(7) | 5(4) |
| Study focus  Epidemiological focus | 76(38) | 3(4) | 5(7) | 3(4) |
| Therapy focus | 34(17) | 0(0) | 0(0) | 10(30) |
| Diagnostic focus | 21(11) | 0(0) | 2(10) | 1(5) |
| Prevention focus | 14(7) | 0(0) | 2(14) | 4(29) |
| Risk focus | 13(7) | 0(0) | 0(0) | 1(8) |
| Pathophysiology focus | 9(5) | 0(0) | 1(11) | 0(0) |
| Other focus | 1(1) | 0(0) | 0(0) | 0(0) |
| **Study focus** N(%) | **Total of subgroup** N(%) | **Code sharing** N(%) | **Data sharing** N(%) | **Registration** N(%) |
| Specific pathogens | 176 (88) | 3(2) | 11(6) | 19(11) |
| Specific disease | 21(11) | 0(0) | 1(5) | 0(0) |
| Specific drug | 36(18) | 0(0) | 1(3) | 11(31) |
| **Study population** | **Total of subgroup** N(%) alt. study median (IQR) | **Code sharing** N(%) alt. study median (IQR) | **Data sharing** N(%) alt. study median (IQR) | **Registration** N(%) alt. study median (IQR) |
| Sample size | 229(1367) | 2159 (1776) | 43(129) | 114(577) |
| Age group  Adult | 96(48) | 1(1) | 3(3) | 10(10) |
| Children | 26(13) | 1(4) | 1(4) | 5(20) |
| Both | 19(10) | 0(0) | 0(0) | 0(0) |
| *Study object*  Humans | 160(80) | 2(1) | 6(4) | 19(12) |
| Pathogen | 16(8) | 0(0) | 3(19) | 0(0) |
| Animal | 6(3) | 0(0) | 2(33) | 0(0) |
| In vitro | 5(3) | 0(0) | 1(20) | 0(0) |
| Vector | 1(1) | 0(0) | 0(0) | 1(100) |
